# Risk factors for severe PCR-positive SARS-CoV-2 infection in hospitalized children: a multicenter cohort study

**DOI:** 10.1101/2021.10.28.21265616

**Authors:** Tilmann Schober, Chelsea Caya, Michelle Barton, Ann Bayliss, Ari Bitnun, Jennifer Bowes, Helena Brenes-Chacon, Jared Bullard, Suzette Cooke, Tammie Dewan, Rachel Dwilow, Tala El Tal, Cheryl Foo, Peter Gill, Behzad Haghighi Aski, Fatima Kakkar, Janell Lautermilch, Ronald M. Laxer, Marie-Astrid Lefebvre, Kirk Leifso, Nicole Le Saux, Alison Lopez, Ali Manafi, Shaun K. Morris, Alireza Nateghian, Luc Panetta, Dara Petel, Dominique Piché, Rupeena Purewal, Lea Restivo, Ashley Roberts, Manish Sadarangani, Rosie Scuccimarri, Alejandra Soriano-Fallas, Sarah Tehseen, Karina A. Top, Rolando Ulloa-Gutierrez, Isabelle Viel-Thériault, Jacqueline K. Wong, Carmen Yea, Ann Yeh, Adriana Yock-Corrales, Joan Robinson, Jesse Papenburg

**Affiliations:** Department of Pediatrics, McGill University, Montreal, Quebec, Canada; Research Institute of the McGill University Health Centre, Montreal, Quebec, Canada; Department of Pediatrics, Western University, London, Ontario, Canada; Department of Pediatrics, Trillium Health Partners, Mississauga, Ontario, Canada; Department of Pediatrics, University of Toronto, Toronto, Ontario, Canada; Department of Pediatrics, University of Ottawa, Ottawa, Ontario, Canada; Department of Pediatrics, Hospital Nacional de Niños “Dr. Carlos Sáenz Herrera”, Caja Costarricense de Seguro Social (CCSS); San José, Costa Rica; Department of Pediatrics, University of Manitoba, Winnipeg, Manitoba, Canada; Department of Pediatrics, University of Calgary, Calgary, Alberta, Canada; Department of Pediatrics, Memorial University, St John’s, Newfoundland and Labrador, Canada; Department of Pediatrics, Iran University of Medical Sciences, Tehran, Iran; Department of Pediatrics, Université de Montréal, Montreal, Quebec, Canada; British Columbia Children’s Hospital, Vancouver, British Columbia, Canada; Department of Pediatrics, University of Saskatchewan, Saskatoon, Saskatchewan, Canada; Department of Pediatrics, Queen’s University, Kingston, Ontario, Canada; Department of Pediatrics, University of British Columbia, Vancouver, British Columbia, Canada; Vaccine Evaluation Center, BC Children’s Hospital Research Institute, Vancouver, British Columbia, Canada; Department of Pediatrics, Dalhousie University, Halifax, Nova Scotia, Canada; Department of Pediatrics, CHU de Québec-Université Laval, Quebec, Quebec, Canada; Department of Pediatrics, McMaster University, Hamilton, Ontario, Canada; Department of Pediatrics, University of Alberta, Edmonton, Alberta, Canada

## Abstract

**Importance:** Children are less likely than adults to have severe outcomes from SARS-CoV-2 infection and the corresponding risk factors are not well established.

**Objective:** To identify risk factors for severe disease in symptomatic children hospitalized for PCR-positive SARS-CoV-2 infection.

**Design:** Cohort study, enrollment from February 1, 2020 until May 31, 2021

**Setting:** 15 children’s hospitals in Canada, Iran, and Costa Rica

**Participants:** Patients <18 years of age hospitalized with symptomatic SARS-CoV-2 infection, including PCR-positive multisystem inflammatory syndrome in children (MIS-C)

**Exposures:** Variables assessed for their association with disease severity included patient demographics, presence of comorbidities, clinical manifestations, laboratory parameters and chest imaging findings.

**Main Outcomes and Measures:** The primary outcome was severe disease defined as a WHO COVID-19 clinical progression scale of ≥6, i.e., requirement of non-invasive ventilation, high flow nasal cannula, mechanical ventilation, vasopressors, or death. Multivariable logistic regression was used to evaluate factors associated with severe disease.

**Results:** We identified 403 hospitalizations. Median age was 3.78 years (IQR 0.53-10.77). At least one comorbidity was present in 46.4% (187/403) and multiple comorbidities in 18.6% (75/403). Severe disease occurred in 33.8% (102/403). In multivariable analyses, presence of multiple comorbidities (adjusted odds ratio 2.24, 95% confidence interval 1.04-4.81), obesity (2.87, 1.19-6.93), neurological disorder (3.22, 1.37-7.56), anemia, and/or hemoglobinopathy (5.88, 1.30-26.46), shortness of breath (4.37, 2.08-9.16), bacterial and/or viral coinfections (2.26, 1.08-4.73), chest imaging compatible with COVID-19 (2.99, 1.51-5.92), neutrophilia (2.60, 1.35-5.02), and MIS-C diagnosis (3.86, 1.56-9.51) were independent risk factors for severity. Comorbidities, especially obesity (40.9% vs 3.9%, p<0.001), were more frequently present in adolescents ≥12 years of age. Neurological disorder (3.16, 1.19-8.43) in children <12 years of age and obesity (3.21, 1.15-8.93) in adolescents were the specific comorbidities associated with disease severity in age-stratified adjusted analyses. Sensitivity analyses excluding the 81 cases with MIS-C did not substantially change the identified risk factors.

**Conclusions and Relevance:** Pediatric risk factors for severe SARS-CoV-2 infection vary according to age and can potentially guide vaccination programs and treatment approaches in children.

**Key points:** *Question:* What are the risk factors for severe disease in children hospitalized for PCR-positive SARS-CoV-2 infection?

*Findings:* In this multinational cohort study of 403 children, multiple comorbidities, obesity, neurological disorder, anemia, and/or hemoglobinopathy, shortness of breath, bacterial and/or viral coinfections, chest imaging compatible with COVID-19, neutrophilia, and MIS-C diagnosis were independent risk factors for severity. The risk profile and presence of comorbidities differed between pediatric age groups, but age itself was not associated with severe outcomes.

*Meaning:* These results can inform targeted treatment approaches and vaccine programs that focus on patient groups with the highest risk of severe outcomes.

## Introduction

The clinical presentation of severe acute respiratory syndrome coronavirus-2 (SARS-CoV-2) infection is generally milder in children than in adults. However, children can also experience morbidity and mortality due to coronavirus disease 2019 (COVID-19).^1^ It is important to identify risk factors for severe disease. This would allow for identification of patient groups that require closer monitoring or early treatment, and could also help guide targeted vaccination campaigns.

While many countries started a vaccination program against SARS-CoV-2 for all adolescents older than 11 years of age in June/July 2021, others had limited access to vaccines or decided to focus on high-risk groups following WHO recommendations.^2,3^ Considering the usually benign clinical course of SARS-CoV-2 infection in children, the potential direct and societal benefits of universal versus targeted vaccination in the pediatric population need to be considered carefully. It is therefore of great practical value to know which children are at high risk of severe disease.

Previous studies have attempted to identify risk factors for severe disease in pediatric patients for severe SARS-CoV-2 infection.^4-14^ Some risk factors such as presence of comorbidities, obesity, and increased inflammatory markers have been implicated,^5,8,10-13,15^ but many studies were small and there is substantial heterogeneity.^16^ Importantly, most major studies were from high-income countries and data from low and middle-income countries are scarce.

Multisystem inflammatory syndrome in children (MIS-C) is a serious post-infectious complication of SARS-CoV-2 infection in children. Despite differences in the pathophysiology and clinical presentation,^17,18^ there is significant overlap between acute SARS-CoV-2 infection and PCR-positive MIS-C; it is frequently impossible to distinguish them and both contribute to the potentially preventable burden of SARS-CoV-2 in children.

The aim of this study was the identification of risk factors for severe disease in children hospitalized for symptomatic PCR-confirmed SARS-CoV-2 infection including PCR-positive MIS-C in a multicenter cohort study from Canada, Costa Rica, and Iran.

## Methods

### Study Design and Population

Thirteen pediatric hospitals from the Canadian Pediatric Investigators Collaborative Network on Infections in Canada (PICNIC) group and two international collaborators (one each in Tehran, Iran, and San José, Costa Rica) entered all consecutive children <18 years of age admitted from February 1, 2020 through May 31, 2021 with PCR confirmed SARS-CoV-2 infection or MIS-C. Cases were identified through the microbiology laboratory, infection prevention and control and/or COVID-19 unit databases. The present study was restricted to symptomatic cases with detection of SARS-CoV-2 by molecular testing from a respiratory specimen during the current admission. Patients who were admitted for reasons other than their SARS-CoV-2 infection (i.e. incidental infections), as per each local investigator’s adjudication, were excluded.

Ethics approval was obtained from each participating institution’s ethics research board, with waiver for informed consent for health records research (Supplementary methods). The STrengthening the Reporting of OBservational studies in Epidemiology (STROBE) guidelines were followed.^19^

### Data Collection

Data on primary indication for admission, demographics, comorbidities, clinical presentation and course, coinfections, treatments, and complications were abstracted by the investigators from the patient’s medical charts and managed using REDCap electronic data capture tools.

### Variables of Interest and Study Definitions

The primary outcome, severe PCR-positive SARS-CoV-2 infection, was defined as a WHO clinical progression scale^20^ of ≥6, i.e. oxygen by non-invasive ventilation or high flow or mechanical ventilation or vasopressors or death. For a secondary analysis, severe SARS-CoV-2 infection was defined as a WHO clinical progression scale^20^ of ≥6 and/or admission to an intensive care unit (ICU). Baseline exposure variables included age, sex country of admission, admission period (arbitrarily divided into prior to July 1, 2020; July 1 until Dec 31 2020; Jan 1 until May 31, 2021), and comorbidities. These chronic health conditions were classified using the Canadian National Advisory Committee on Immunization risk group classification for influenza^21^ and adapted according to putative risk factors for severe COVID-19 in children (Supplementary methods). Other potential predictor variables (measured at any point during the hospitalization) were: clinical manifestations, viral co-infection (defined as laboratory detection of any other virus; testing at the discretion of the clinical team), bacterial co-infection (defined as laboratory detection of ≥1 bacteria treated with antibiotics; testing and treatment at the discretion of the clinical team), chest imaging abnormalities (chest x-ray or CT compatibility with COVID-19 based on the reporting physician’s assessment), laboratory parameters (age-defined leukocytosis, leukopenia, neutrophilia, neutropenia, thrombocytosis or thrombocytopenia^22^; C-reactive Protein [CRP] >50mg/L; Ferritin >5000 mcg/L; albumin <29g/L), diagnosis of MIS-C (as defined by WHO criteria^23^).

### Data Analysis

Data were analyzed using R version 3.5.2 (R Core Team, Vienna, Austria). Statistical significance was assessed by using 2-tailed tests, with an α of 0.05. Baseline and demographic characteristics were summarized using descriptive statistics. Categorical data were compared using chi-square or Fisher’s exact test as appropriate and continuous data compared using Wilcoxon rank sum test. Statistical testing was limited to hypothesis-driven and biologically plausible comparisons; accordingly, correction for multiple hypothesis testing was not done. Logistic regression analyses were performed to determine crude and adjusted odds ratios (aORs) and corresponding 95% confidence intervals (CIs) for factors associated with se1vere disease. For the multivariable models, we considered variables with p-value ≤0.2 on univariable analyses and, a priori, variables which were potential confounders based on literature or clinical experience. Model selection was guided by Akaike Information Criteria, using backward and forward selection. Pre-specified sensitivity (excluding MIS-C cases) and age-stratified analyses (children under 12 years old and adolescents ≥ 12 years old) were also performed.

## Results

From February 1, 2020 through May 31, 2021, 982 children younger than 18 years of age hospitalized for SARS-CoV-2-related illness and/or PCR-positive MIS-C were reported from the 15 study hospitals. We excluded 579 cases in whom SARS-CoV-2 was not confirmed by molecular testing from respiratory specimens or because the patient was admitted for non-SARS-CoV-2-related reasons. We included the remaining 403 hospitalizations (Figure 1).

**Figure 1.**
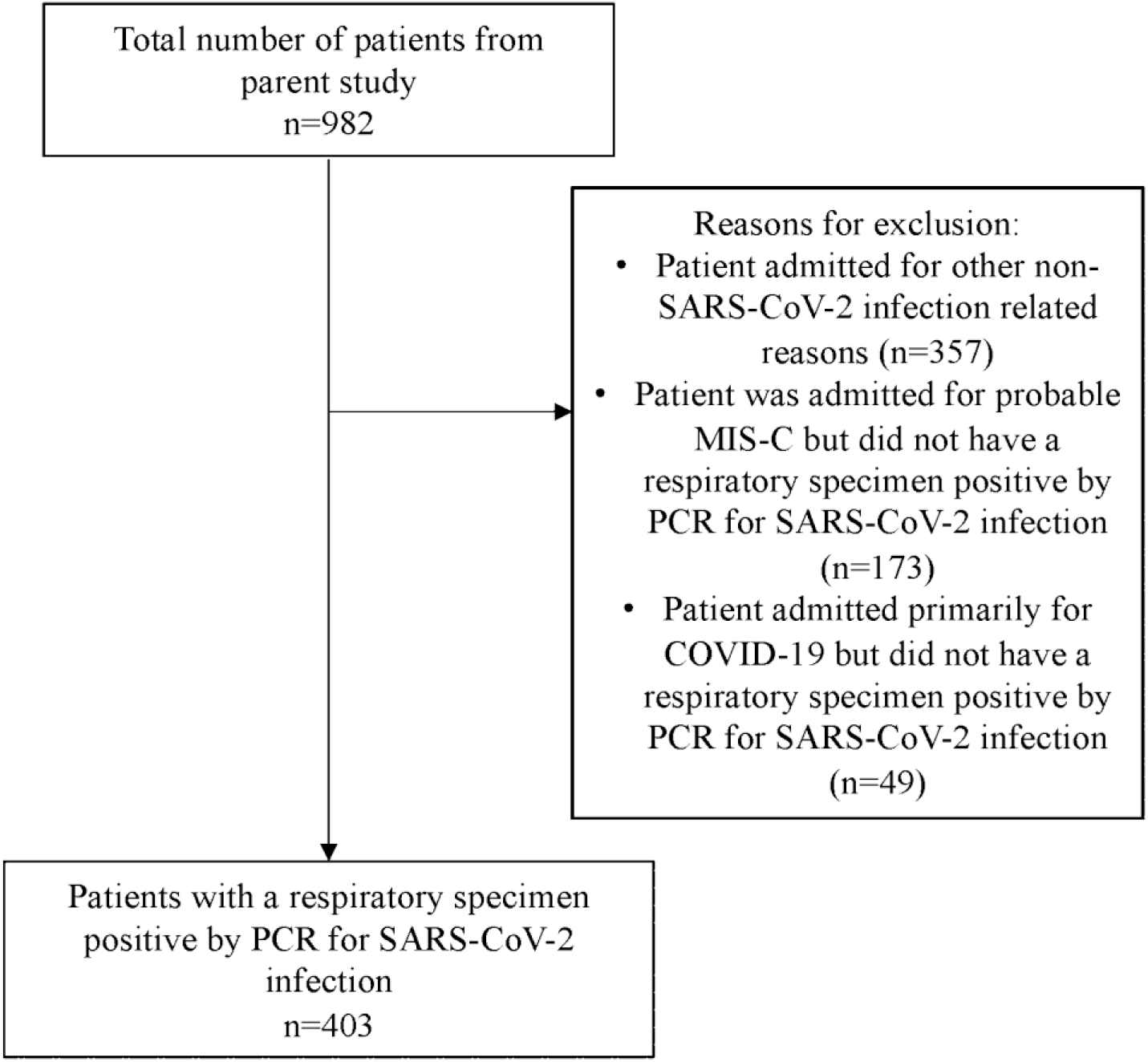
Flow chart of hospitalized children <18 years of age reported to the PICNIC study between February 1, 2020 and May 31, 2021 with a diagnosis of SARS-CoV-2 infection

### Demographics and Clinical Characteristics

Hospitalizations occurred from March 19^th^ 2020 through May 31^st^ 2021; 266 cases (66%) were from Canada, 107 (27%) from Costa Rica, and 30 (7%) from Iran (Tables 1 and 2). Overall median age was 3.78 years (interquartile range [IQR]: 0.53-10.77). At least 1 comorbidity was present in 46.4% and multiple comorbidities in 18.6% of children. Fever (82.1%), cough (51.6%), shortness of breath (46.7%), vomiting (31.5%), rhinitis (31%), diarrhea (28.5%), and abdominal pain (26.8%) were the most common presenting symptoms. Intensive care unit (ICU) admission was required in 115 patients (28.5%). Six patients died, all with comorbidities, including 2 patients with malignancies receiving palliative care.

**Table 1.**
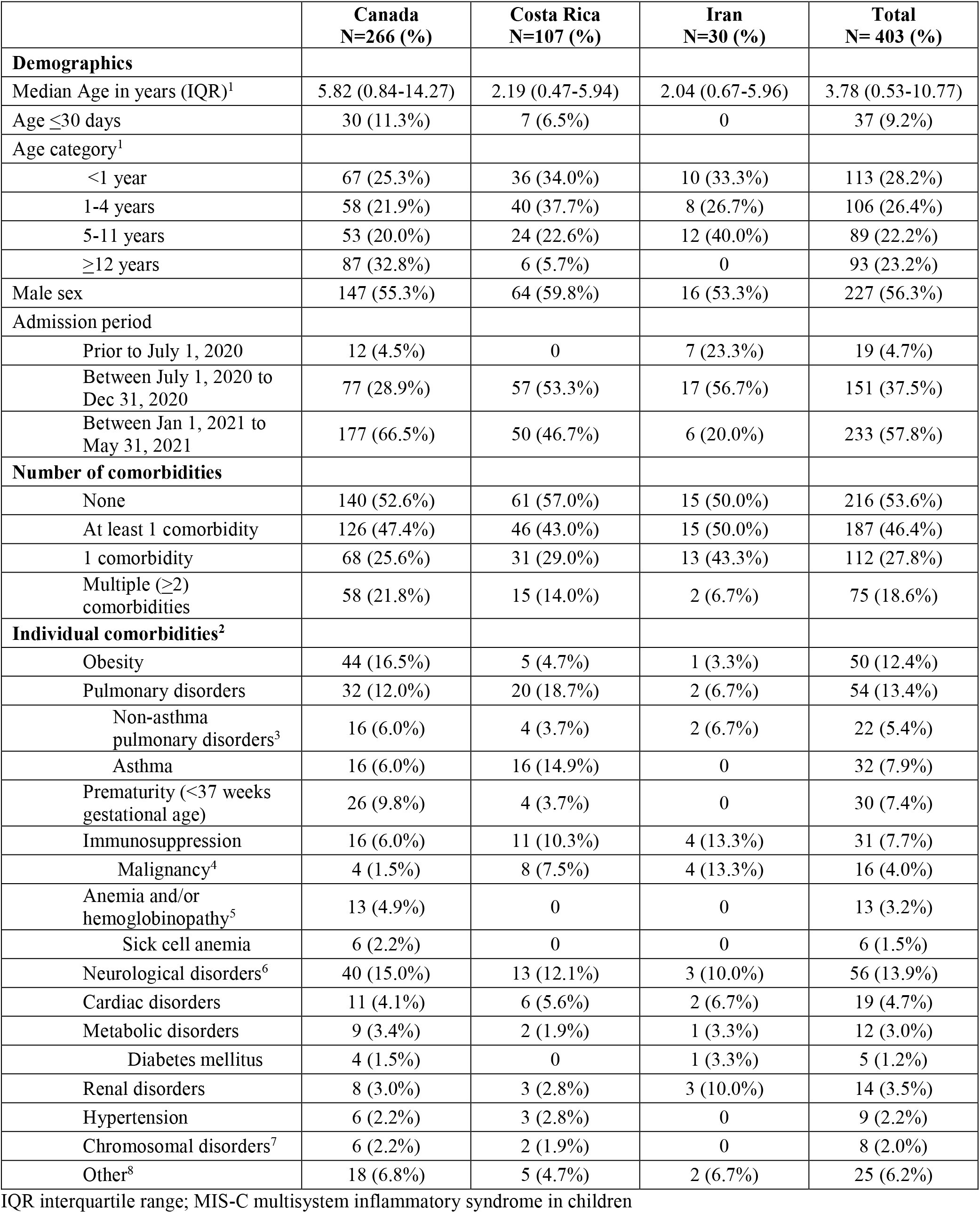
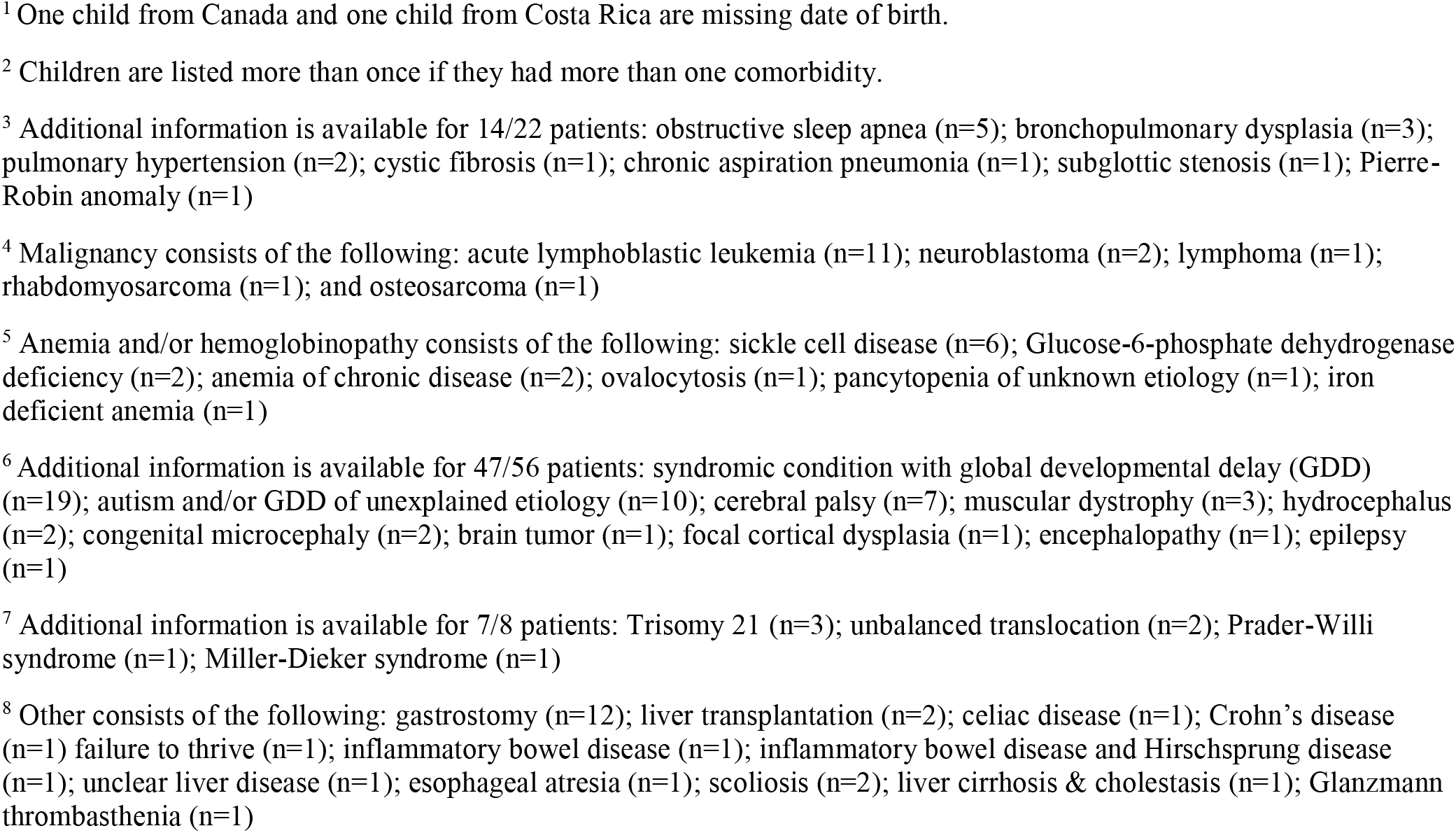
Baseline characteristics of patients with PCR-positive SARS-CoV-2 infection (n=403) according to country of origin

**Table 2.** Clinical characteristics, investigations, treatments and outcomes of patients with severe PCR-positive SARS-CoV-2 infection (n=403) according to country of origin

|  | **Canada**<br>**N=266 (%)** | **Costa Rica**<br>**N=107 (%)** | **Iran**<br>**N=30 (%)** | **Total**<br>**N= 403 (%)** |
| --- | --- | --- | --- | --- |
| **Clinical Features** |  |  |  |  |
| Fever prior to or during admission<sup>1</sup> | 205 (82.3%) | 71 (78.0%) | 26 (92.8%) | 302 (82.1%) |
| Cough | 136 (51.1%) | 54 (50.5%) | 18 (60.0%) | 208 (51.6%) |
| Shortness of breath | 125 (47.0%) | 55 (51.4%) | 8 (26.7%) | 188 (46.7%) |
| Vomiting | 91 (34.2%) | 22 (20.6%) | 14 (46.7%) | 127 (31.5%) |
| Rhinitis | 66 (24.8%) | 49 (45.8%) | 10 (33.3%) | 125 (31.0%) |
| Diarrhea | 70 (26.3%) | 31 (29.0%) | 14 (46.7%) | 115 (28.5%) |
| Abdominal pain | 67 (25.2%) | 26 (24.3%) | 15 (50.0%) | 108 (26.8%) |
| Headache | 47 (17.7%) | 19 (17.8%) | 6 (20.0%) | 72 (17.9%) |
| Rash | 42 (15.8%) | 24 (22.4%) | 3 (10.0%) | 69 (17.1%) |
| Conjunctivitis | 46 (17.3%) | 21 (19.6%) | 1 (3.3%) | 68 (16.9%) |
| Myalgia | 30 (11.3%) | 12 (11.2%) | 18 (60.0%) | 60 (14.9%) |
| Pharyngitis | 32 (12.0%) | 15 (14.0%) | 13 (43.3%) | 60 (14.9%) |
| Wheeze | 16 (6.0%) | 26 (24.3%) | 12 (40.0%) | 54 (13.4%) |
| Chest pain | 32 (12.0%) | 3 (2.8%) | 4 (13.3%) | 39 (9.7%) |
| Cracked lips | 22 (8.3%) | 16 (14.9%) | 0 | 38 (9.4%) |
| Edema of hands and feet | 14 (5.3%) | 12 (11.2%) | 1 (3.3%) | 27 (6.7%) |
| Anosmia | 10 (3.7%) | 2 (1.9%) | 0 | 12 (3.0%) |
| Seizures | 6 (2.2%) | 3 (2.8%) | 2 (6.7%) | 11 (2.7%) |
| **Coinfections** |  |  |  |  |
| Bacterial (positive cultures)<sup>2</sup> | 14 (5.3%) | 7 (6.5%) | 3 (10.0%) | 24 (5.9%) |
| Viral | 7 (2.6%) | 25 (23.4%) | 0 | 32 (7.9%) |
| Bacterial and/or viral | 21 (7.9%) | 30 (28.0%) | 3 (10.0%) | 54 (13.4%) |
| **Investigations** |  |  |  |  |
| Chest imaging performed<sup>3</sup> | 200 (75.2%) | 82 (76.6%) | 30 (100.0%) | 312 (77.4%) |
| CXR and/or CT chest normal or had findings unrelated to COVID-19 | 60 (22.5%) | 45 (42.0%) | 16 (53.3%) | 121 (30.0%) |
| CXR and/or CT chest abnormalities compatible with COVID-19 | 140 (52.6%) | 37 (34.6%) | 14 (46.7%) | 191 (47.4%) |
| **Laboratory investigations<sup>4</sup>** |  |  |  |  |
| Leukopenia (<4 X10<sup>9</sup>/L)<sup>5</sup> | 43 (17.1%) | 12 (11.5%) | 4 (13.3%) | 59 (15.3%) |
| Leukocytosis (>15x10<sup>9</sup>/L)<sup>5</sup> | 20 (7.9%) | 21 (20.2%) | 6 (20.0%) | 47 (12.2%) |
| Neutropenia (<1.5x 10<sup>9</sup>/L)<sup>6</sup> | 53 (21.4%) | 27 (26.0%) | 8 (26.7%) | 88 (23.0%) |
| Neutrophilia<sup>6</sup> | 73 (29.4%) | 37 (35.6%) | 10 (33.3%) | 120 (31.4%) |
| Thrombocytopenia (<100 x 10<sup>9</sup>/L)<sup>7</sup> | 27 (10.8%) | 12 (11.5%) | 5 (16.7%) | 44 (11.4%) |
| Thrombocytosis (>450 x 10<sup>9</sup>/L)<sup>7</sup> | 47 (18.7%) | 29 (27.9%) | 10 (33.3%) | 86 (22.3%) |
| CRP >50 (mg/L)<sup>8</sup> | 87 (38.7%) | 22 (25.0%) | 10 (33.3%) | 119 (34.7%) |
| Ferritin >500 (mcg/L)<sup>9</sup> | 37 (28.2%) | 12 (37.5%) | 6 (30.0%) | 55 (30.0%) |
| Albumin<29 (g/L)<sup>10</sup> | 34 (45.9%) | 10 (21.7%) | 2 (16.7%) | 46 (34.8%) |
| **Treatments** |  |  |  |  |
| Intravenous immunoglobulin | 48 (18.0%) | 22 (20.6%) | 12 (40.0%) | 82 (20.3%) |
| Antiviral agent for influenza | 4 (1.5%) | 0 | 4 (13.3%) | 8 (2.0%) |
| Corticosteroids | 130 (48.9%) | 30 (28.0%) | 8 (26.7%) | 168 (41.7%) |
| Lopinavir-ritonavir | 0 | 0 | 8 (26.7%) | 8 (2.0%) |
| Remdesivir | 35 (13.1%) | 0 | 6 (20.0%) | 41 (10.2%) |
| Azithromycin | 0 | 0 | 13 (43.3%) | 13 (3.2%) |
| Hydroxychloroquine | 1 (0.4%) | 0 | 23 (76.7%) | 24 (5.9%) |
| **Highest level of care required** |  |  |  |  |
| No supplemental oxygen or ICU admission | 137 (51.5%) | 38 (35.5%) | 18 (60.0%) | 193 (47.9%) |
| Supplemental oxygen but no ICU admission<sup>11</sup> | 47 (17.7%) | 46 (43.0%) | 2 (6.7%) | 95 (23.6%) |
| ICU admission | 82 (30.8%) | 23 (21.5%) | 10 (33.3%) | 115 (28.5%) |
| ICU care without CPAP/BIPAP/ MV | 45 (16.9%) | 11 (10.3%) | 6 (20.0%) | 62 (15.4%) |
| ICU with CPAP/BIPAP | 18 (6.8%) | 2 (1.9%) | 0 | 20 (5.0%) |
| ICU with MV | 19 (7.1%) | 10 (9.3%) | 4 (13.3%) | 33 (8.2%) |
| Extracorporeal life support | 0 | 0 | 0 | 0 |
| **Complications** |  |  |  |  |
| MIS-C diagnosis | 59 (22.2) | 19 (17.7) | 3 (10.0) | 81 (20.1%) |
| **Outcomes** |  |  |  |  |
| Mean length of hospitalization in days (SD)<sup>12</sup> | 5.9 (6.0) | 6.5 (8.1) | 9.3 (6.6) | 6.3 (6.7) |
| Median length of hospitalization in days (IQR) | 4.0 (2.0-7.0) | 4.0 (3.0-7.0) | 7.0 (6.0-9.0) | 4.0 (2.0-7.0) |
| Death | 2 (0.7%) | 3 (2.8%) | 1 (3.3%) | 6 (1.5%) |
| Severe disease according to WHO definition | 71 (26.7%) | 27 (25.2%) | 4 (13.3%) | 102 (25.3%) |
| Severe disease according to WHO definition and/or ICU admission | 91 (34.2%) | 27 (25.2%) | 10 (33.3%) | 128 (31.8%) |

BiPAP bilevel positive airway pressure; CPAP continuous positive airway pressure; CT computed tomography; CRP C-reactive protein; CXR chest x-ray; ICU intensive care unit; IQR interquartile range; MIS-C multisystem inflammatory syndrome in children; MV mechanical ventilation; SD standard deviation
<sup>1</sup> A total of 35 children were missing data on fever prior to or during admission (17 from Canada, 16 from Costa Rica and 2 from Iran) resulting in a total N of 368.
<sup>2</sup> Children with one of the following are included: blood culture thought to be truly positive and/or positive endotracheal tube cultures treated with antibiotics for presumptive bacterial pneumonia and/or positive bacterial culture from another site treated with antibiotics.
<sup>3</sup> Children are listed more than once if they had more than one chest imaging result (e.g., chest x-ray and or chest CT).
<sup>5</sup> A total of 17 children were missing leukocyte values (14 from Canada and 3 from Costa Rica) resulting in a total N of 386.
<sup>6</sup> A total of 21 children were missing neutrophil values (18 from Canada and 3 from Costa Rica) resulting in a total N of 382. Cut-off values for neutrophilia are age dependent.
<sup>7</sup> A total of 18 children were missing platelet values (15 from Canada and 3 from Costa Rica) resulting in a total N of 385.
<sup>8</sup> A total of 60 children were missing a CRP value (41 from Canada and 19 from Costa Rica) resulting in a total N of 343.
<sup>9</sup> The variable assessing highest ferritin value used and a total of 220 children were missing a value (135 from Canada, 75 from Costa Rica and 10 from Iran) resulting in a total N of 183.

Compared to the two middle-income countries, cases from Canada were older (median 5.82 [IQR 0.84-14.27] vs 2.15 [IQR: 0.48-6.01] years) with a higher proportion of adolescents (32.8% vs 5.6%, p<0.001) and occurred later during the study period (66.5% vs 40.8% admitted in 2021, p<0.001). Other key demographic, clinical and outcome parameters were similar between countries, including length of hospitalization, proportion requiring ICU admissions, severity according to WHO definition and percentage of cases with MIS-C.

### Predictors of Disease Severity

The primary outcome of severe disease (WHO COVID-19 clinical progression scale ≥6) occurred in 102 (25%) of 403 children (Table 3). In univariable analysis, patients with severe disease were older (median 6.69 [IQR 1.31-13.46] vs 3.21 [IQR 0.31-9.87] years, p=0.002). The proportion of children with severe disease increased progressively with age, with the lowest prevalence in neonates (8.1%) and the highest in ≥12 year olds (35.5%). Compared with previously healthy patients, those with 1 or with ≥2 comorbidities were more likely to have severe disease (19.9%, 31.5%, and 37.3% respectively; p=0.009). Among individual comorbidities, obesity, non-asthma pulmonary disease, hypertension, neurological and chromosomal disorders were also risk factors, but not immunosuppression (Table 3). Shortness of breath was the only clinical presentation associated with severity with 41.5% having severe disease. Among laboratory and imaging investigations, viral coinfection, chest imaging compatible with COVID-19, neutrophilia, increased CRP, and low albumin predicted a severe course. In contrast, bacterial coinfections, abnormal leukocyte or platelet count, and increased ferritin were not statistically different between mild/moderate and severe cases.

**Table 3.**
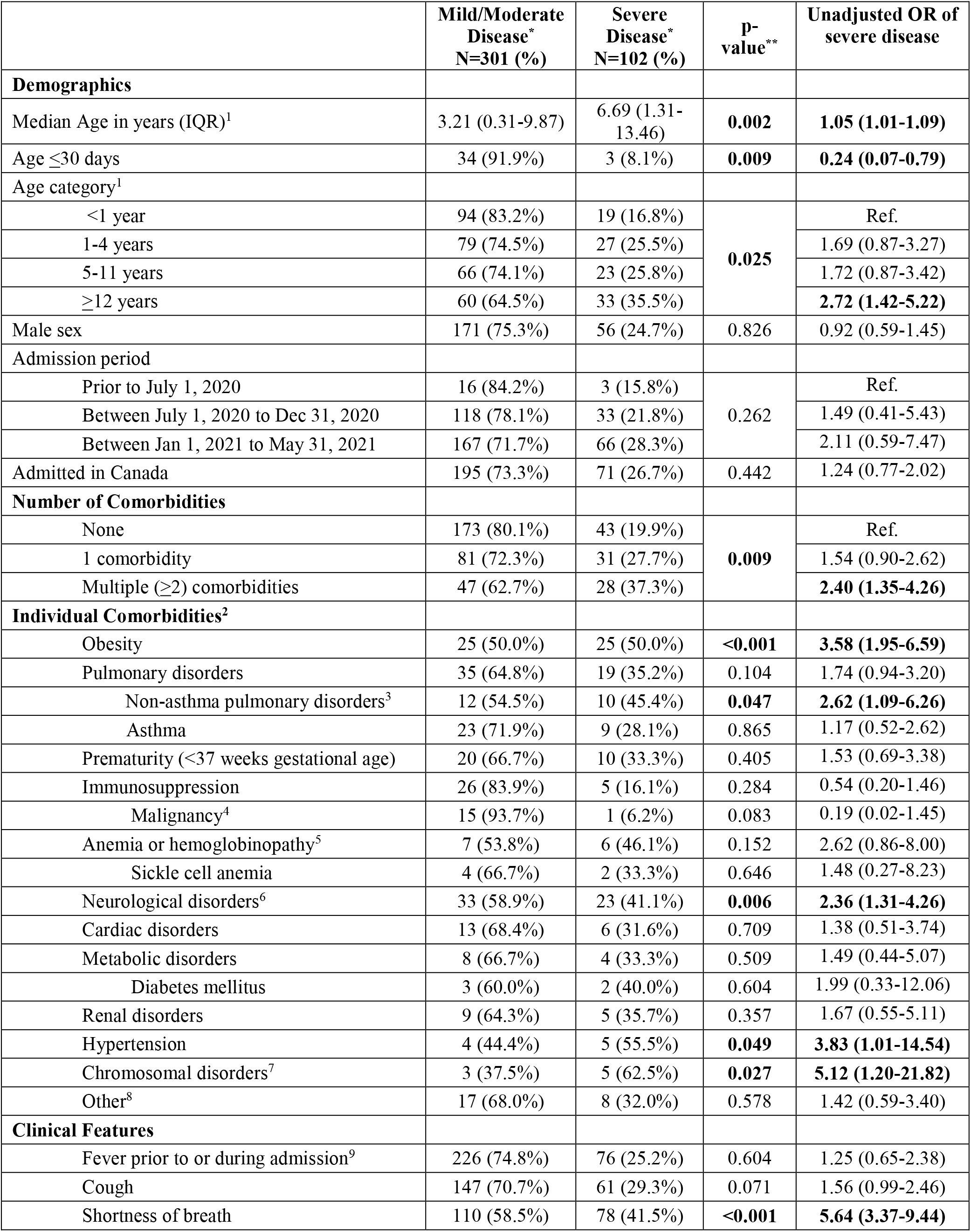
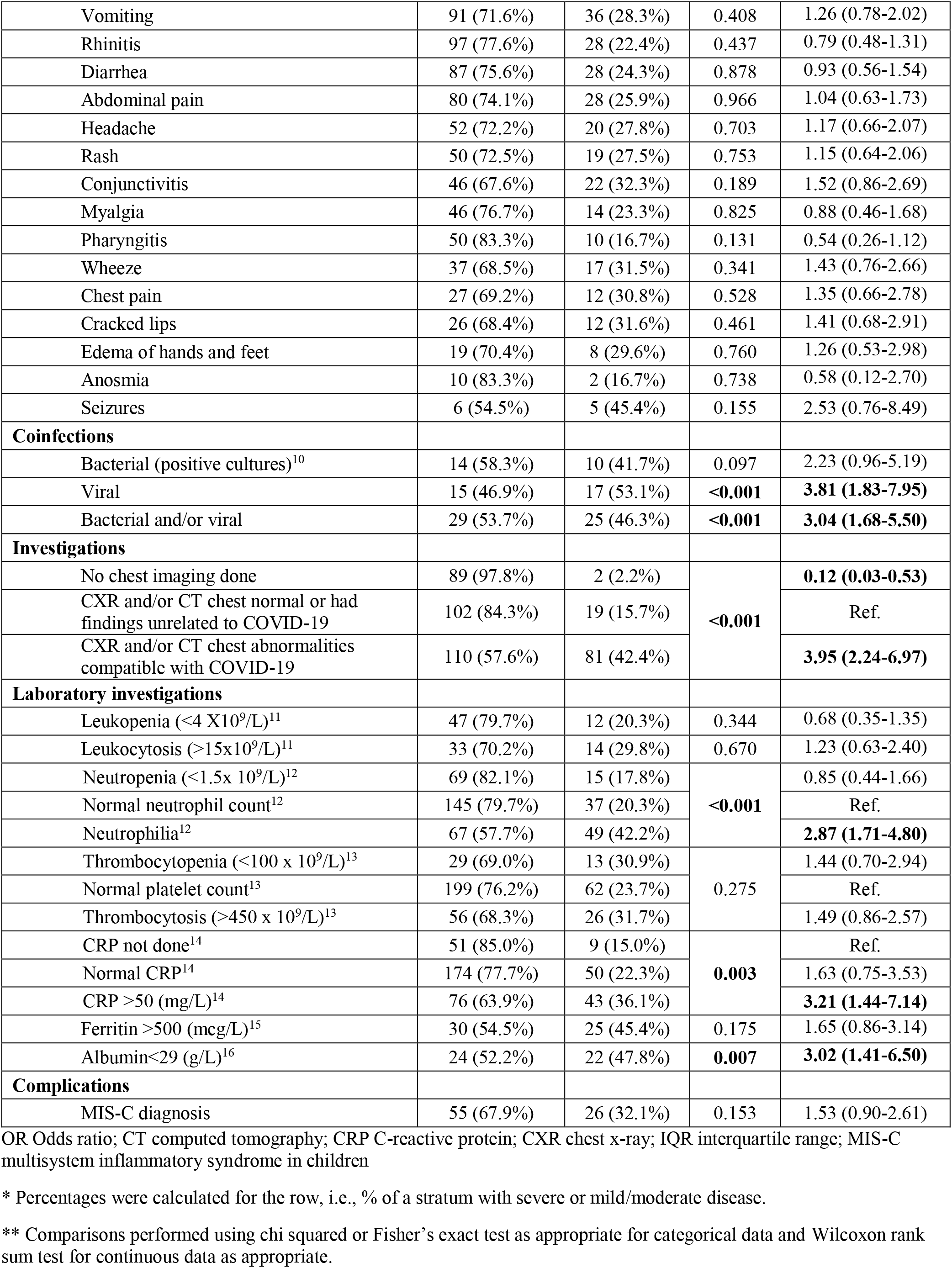
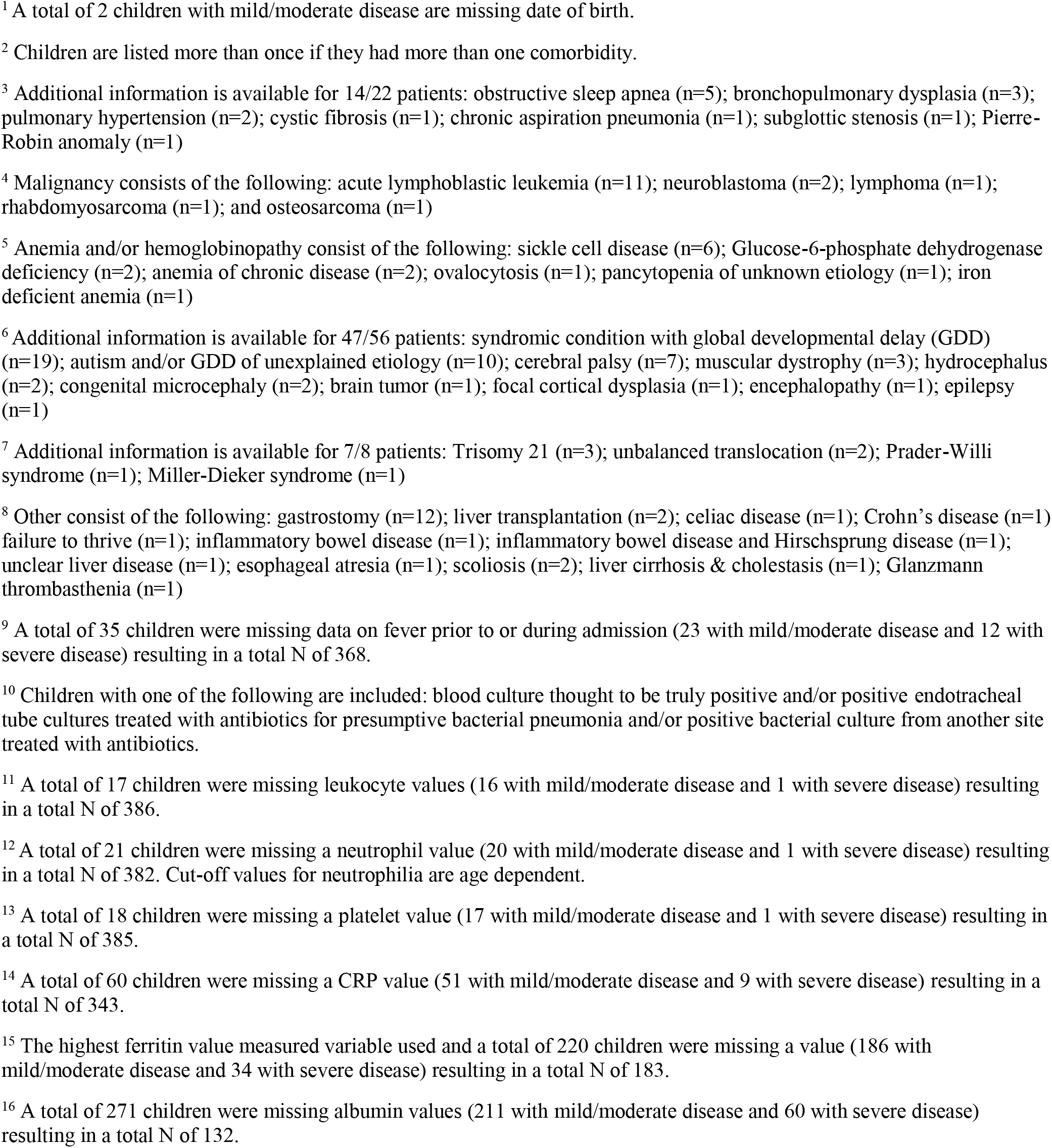
Characteristics of patients with severe PCR-positive SARS-CoV-2 infection (n=403) according to severity of disease (WHO COVID-19 clinical progression scale of ≥6).

Multivariable logistic regression analysis demonstrated that obesity (OR 2.87, 95% CI 1.19-6.93), pre-existing anemia and/or hemoglobinopathy (OR 5.88, 95% CI 1.30-26.46), underlying neurological disorder (OR 3.22, 95% CI 1.37-7.56), shortness of breath (OR 4.37, 95% CI 2.08-9.16), bacterial and/or viral coinfections (OR 2.26, 95% CI 1.08-4.73), chest imaging compatible with COVID-19 (OR 2.99, 95% CI 1.51-5.92), neutrophilia (OR 2.60, 95% CI 1.35-5.02), and MIS-C (OR 3.86, 95% CI 1.56-9.51) diagnosis were independent predictors of severe disease (Figure 2). In a separate model adjusting for the same covariates, we examined the effect of the number of comorbidities instead of each comorbidity individually. The presence of multiple comorbidities (OR 2.24, 95% CI 1.04-4.81), but not of a single comorbidity (OR 1.71, 95% CI 0.86-3.43) was a statistically significant risk factor (Figure 2). Limiting the multivariable analysis to baseline characteristics identified the same risk factors as the aforementioned models which included clinical, laboratory, and outcome parameters (eTable 1).

**Figure 2.**
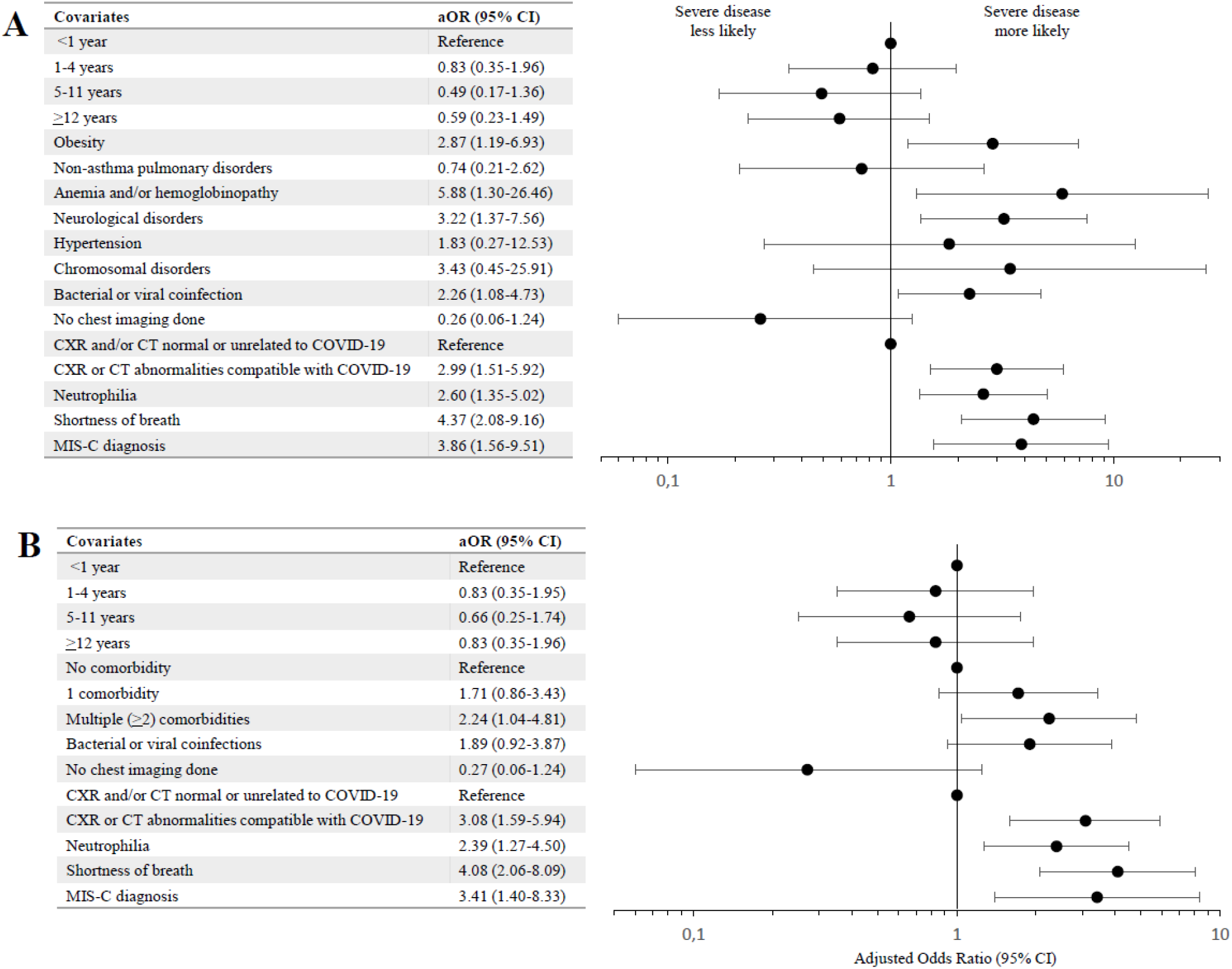
Multivariable logistic regression models for factors associated with severe PCR-positive SARS-CoV-2 infection (WHO COVID-19 clinical progression scale of ≥6) **Legend**: The model in Panel A evaluated individual comorbidities whereas the model in Panel B evaluated the number of comorbidities, as risk factors for disease severity. aOR adjusted odds ratio; CT computed tomography; CXR chest x-ray; MIS-C multisystem inflammatory syndrome in children

We further analyzed the data using the secondary outcome of ICU admission and/or a WHO COVID-19 clinical progression scale of ≥6. There were 128 severe cases (31.8%) using this definition. ICU admission as a sole criterion excluded 10 patients with high flow nasal cannula and/or non-invasive ventilation who were managed outside of an ICU. Of 118 patients admitted to ICU, 26 (22%) did not meet our primary severity definition. Contrary to our primary unadjusted analyses, single comorbidity, chromosomal disorders, and hypertension were not significantly associated with severe disease (eTable 2).

In unadjusted stratified analyses by age, risk factors for severity in children <12 years were nearly identical to the overall study population (eTable 3-4). However, in multivariable models, the only individual comorbidity to remain significantly associated with severity was neurologic disorders (eTable 6). Among adolescents ≥ 12 years of age, the only comorbidity associated with severity was obesity (eTable 6). Of note, risk factors were not equally distributed between the two age groups. More adolescents had any (66.7% vs. 40.2%, p<0.001) or multiple (25.8% vs. 16.5%, p=0.049) comorbidities than younger children; obesity was markedly more common in adolescents (40.9% vs 3.9%, p<0.001).

In a sensitivity analysis, we excluded the 81 cases who met WHO MIS-C criteria. Results were very similar to the overall patient population, except that anemia and/or hemoglobinopathy and neutrophilia lost significance in the multivariable analysis (eTables 7-8).

## Discussion

In this multicenter cohort study from three countries, we identified risk factors for severe disease among children admitted to hospital for PCR-positive symptomatic SARS-CoV-2 infection. Previous studies have been either limited to administrative data,^4,8,12^ were smaller,^5-7,10,11,13,14^ or had a different scope.^18^ We confirmed several previously described risk factors for severity on multivariable analyses: presence of multiple comorbidities,^4,6,9,12-15^ obesity,^5,8,11,14^ neurological disorders,^8,14^ anemia and/or hemoglobinopathy,^24^ shortness of breath,^6^ bacterial and/or viral coinfections,^6,15^ chest imaging compatible with COVID-19^5,13^, neutrophilia,^13^ and MIS-C diagnosis.^5,18^ Additionally, older age,^4,10-12^ hypertension,^8^ and chromosomal disorders,^25^ increased CRP,^10,11,13,15,24^ and low albumin were associated with higher odds of a severe course in unadjusted analyses only, but not in multivariable models, indicating that other factors may have been confounding this association or that our study was underpowered to assess less frequent comorbidities. We also found evidence that age modifies the effect of individual comorbidities, with neurological disorders strongly associated with severe disease in children under 12 years old, whereas only obesity predicted severe disease in adolescents.

Less than 5% of severe cases in our cohort were immunocompromised and impaired immunity was not associated with severity, in keeping with past pediatric studies.^5,13,14^ A recent study on children with cancer showed a higher rate of complications following SARS-CoV-2 infection among the severely immunocompromised.^26^ However, the study lacked a control group and immunocompromised children are extremely heterogeneous with multiple comorbidities complicating the interpretation. Accordingly, our data support the concept that a dysregulated rather than an insufficient immune response is a greater contributor to severe SARS-CoV-2 infection in the majority of children and adults requiring hospitalization.^17^

Previous studies have described a bimodal age pattern regarding severe disease, with a first peak in the very young and a second increase in adolescence mimicking global non-SARS-CoV-2-related childhood mortality.^4,5,7,12^ Infants in our study population were over-represented compared to the general pediatric population. For Canadian children, the proportion of infants was 4.3 times higher than its share of the population,^27^ suggesting a higher risk of hospitalization. It is likely that some young infants with mild symptoms were admitted for observation, especially early during the pandemic, or to rule-out serious bacterial infection.^28^ We did not see a higher rate of severe courses according to age, in line with other studies which restricted their analysis to hospitalized children.^10,14,15^ Infants typically have a significantly higher baseline all-cause morbidity and mortality than older children.^29^ Analyses which include all SARS-CoV-2 positive patients independent of their symptoms may mimic the baseline risk and not necessarily the infection-related complications.

Similar to other studies, we saw a higher rate of severe disease in adolescents in our univariable analysis (Table 3),^4,10-12^ but this was not confirmed in the multivariable models (Figure 2). This age group had a different risk profile than younger children, with obesity as the only comorbidity significantly associated with illness severity. Guzman *et al*. also observed that obesity was an independent risk factor for critical illness in adolescents but not younger children.^30^ This association more closely resembles adults, in whom metabolic risk factors are among the most important.^31^ The observed higher proportion of severe cases among adolescents in crude analyses could be mainly due to the markedly higher prevalence of obesity (40.9% in adolescents vs 3.9% in younger children in our study) and other comorbidities.

Our primary outcome differed from many previous studies which defined severity as ICU admission.^5-8,15,24^ ICU admissions are simple to measure, but can be problematic as a severity outcome because of heterogeneity across study sites due to dependence on local health care delivery structures and practice.

In addition, patients with some comorbidities or within certain age categories may be admitted to ICU mainly for monitoring or for treatment of a condition independent of the severity of the current illness, e.g. patients with tracheostomy or type-1 diabetes mellitus.^32^ Therefore, we used the more objective WHO COVID-19 clinical progression scale which is based on the need for mechanical or medical support of vital functions,^20^ similar to the definition used by Ouldali et al.^10^ Our comparative analysis of two different severity criteria shows that the effect of several important risk factors would have been obscured using ICU admission alone to define severity.

We decided to include all hospitalized symptomatic patients with positive SARS-CoV-2 PCR, independent of the diagnosis of MIS-C. Although the pathophysiology is different,^17^ it is frequently impossible to distinguish severe acute SARS-CoV-2 infections from PCR-positive MIS-C and both can be present at the same time.^18^ In a sensitivity analysis, risk factors for a severe course after exclusion of MIS-C cases were very similar to the primary analysis that included MIS-C. Thus, the strict separation of acute COVID-19 and early PCR-positive MIS-C may be an oversimplification. Moreover, treatment regimens for both conditions can overlap. Late, and typically PCR-negative, MIS-C has been shown to have different severity risk factors.^5,13,18^ By requiring patients to be PCR-positive, two-thirds of MIS-C patients were excluded. The analysis of all MIS-C patients from the PICNIC cohort is being reported separately.^33^

Our study has limitations. First, reporting may not have been uniform across centers despite the use of a study protocol and a standardized data capture tool. This includes but is not limited to the interpretation of chest radiographs, definition of obesity, and the severity of comorbidities. Also, the respective comorbidities can represent a heterogeneous group of diseases with different etiologies. Second, despite being one of the largest pediatric cohort studies with more than 100 severe cases, the numbers of some individual comorbidities are small. This might overestimate or underestimate the impact of infrequent comorbidities such as anemia and/or hemoglobinopathy, chromosomal, or cardiac disorders. Third, laboratory investigations were not systematically performed. Complete blood counts, CRP, and albumin values were missing in 4.2%, 15%, and 67%, respectively. We therefore could not assess the latter in multivariable models. Moreover, laboratory investigations were done at the clinicians’ discretion, not at set time points during the illness. Other parameters previously described as risk factors could not be evaluated. This includes ethnicity^4,7,9,12^ which was not readily accessible from hospital records. In addition, viral sequencing data was not available, accordingly we cannot evaluate the impact of different SARS-CoV-2 variants. Fourth, indications for testing for SARS-CoV-2 likely varied by center and over time, which might have affected the clinical spectrum of included patients. However, since we analyzed symptomatic hospitalized patients admitted for SARS-CoV-2 infection or PCR-positive MIS-C in centers with availability of SARS-CoV-2 PCR tests, we do not expect this to bias our results. Fifth, our study was not population-based. Most participating hospitals are referral centers so milder hospitalized cases might have been missed and this might overestimate the rate of severe courses. In addition, there were only two hospitals from Costa Rica and Iran, representing one-third of the total sample size; they may not be representative of middle-income countries in general.

A major strength of our study includes its multicenter, multi-national design covering the first 15 months of the COVID-19 pandemic. The patient population of symptomatic children hospitalized for PCR-confirmed SARS-CoV-2 is well-defined and reduces potential biases from including incidental detections of SARS-CoV-2. Finally, this study represents one of the largest yet comprehensive datasets with over 100 severe cases, thereby allowing us to identify differences in risk factor profiles across age groups.

In conclusion, we identified several independent risk factors for severe disease in children hospitalized for symptomatic PCR-positive SARS-CoV-2 infection, namely the presence of multiple comorbidities, obesity, anemia and/or hemoglobinopathy, neurological disorder, shortness of breath, bacterial and/or viral coinfections, chest imaging compatible with COVID-19, neutrophilia, and MIS-C diagnosis. Based on our analysis, age was not an independent risk factor, but the apparent increased risk in adolescents is mainly secondary to the presence of age-specific comorbidities. Furthermore, age appears to modify the effect of specific comorbidities such as obesity. Improved understanding of pediatric risk factors for severe SARS-CoV-2 infection among children requiring hospitalization can guide management and refine risk-benefit analyses of pediatric vaccination programs against SARS-CoV-2.

## Acknowledgements

We thank Dr. Joanna Merckx for her thoughtful review of an earlier version of this manuscript.

## Author contributions

The study was conceived by Joan Robinson, Michelle Barton-Forbes and Jesse Papenburg, and supervised by Joan Robinson and Jesse Papenburg. Analysis was conducted by Chelsea Caya and Tilmann Schober. Tilmann Schober wrote the first draft of the manuscript. All authors contributed to data collection and interpretation, reviewed the manuscript critically for important intellectual content, gave final approval of the version to be published and agreed to be accountable for all aspects of the work.

## Data sharing statement

The corresponding authors will consider data sharing requests that are accompanied by a study protocol and approval by an independent research ethics board.

