## Supplement for "Risk factors for severe PCR-positive SARS-CoV-2 infection in hospitalized children: a multicenter cohort study"

### Supplementary Methods

**eTable 1.** Multivariable logistic regression models focusing only on baseline characteristics as risk factors for severe PCR-positive SARS-CoV-2 infection (WHO COVID-19 clinical progression scale of ≥6)

**eTable 2**. Demographic and clinical characteristics of patients PCR-positive SARS-CoV-2 infection (n=403) according to severity of disease (WHO COVID-19 clinical progression scale of ≥6 and/or ICU admission)

**eTable 3** Age stratified univariable logistic regression analysis among children aged younger than 12 years (n=308) and adolescents 12 years of age and older (n=93) for severe PCR-positive SARS-CoV-2 infection (WHO COVID-19 clinical progression scale of ≥6)

**eTable 4.** Demographic and clinical characteristics among children aged less than 12 years (n=308) according to according to severity of disease (WHO COVID-19 clinical progression scale of ≥6)

**eTable 5.** Demographic and clinical characteristics among adolescents aged 12 to 17 years (n=93) according severity of disease (WHO COVID-19 clinical progression scale of ≥6)

**eTable 6.** Multivariable logistic regression models for factors associated with severe PCR-positive SARS-CoV-2 infection (WHO COVID-19 progression scale of ≥6), stratified according to age groups

**eTable 7.** Demographic and clinical characteristics among non-MIS-C patients (n=322) with PCR-positive SARS-CoV-2 infection, according to severity of disease (WHO COVID-19 clinical progression scale of ≥6)

**eTable 8.** Multivariable logistic regression models among non-MIS-C patients for factors associated with severe PCR-positive SARS-CoV-2 infection (WHO COVID-19 clinical progression scale of ≥6).

**Supplementary Methods**

Ethics approval was obtained from the following ethics review boards: Comité Ético Científico Hospital Nacional de Niños, San José, Costa Rica (CEC-HNN-030-2020), Iran University of Medical Sciences Ethics Review Committee (IR.IUMS.REC.1399.187), The Hospital for Sick Children Research Ethics Board (#1000070091), Pediatric Panel of the Research Ethics Board of the Research Institute of the McGill University Health Centre (#MP-37-2021-6561), Conjoint Health Research Ethics Board, University of Calgary (REB20-0594), Children's Hospital of Eastern Ontario Research Ethics Board (CHEOREB# 20/32X), University of British Columbia Children's and Women's Research Ethics Board (# H20-00977), Health Research Ethics Board, University of Manitoba (HSC23858), University of Saskatchewan Biomedical Research Ethics Board (Study 1921), Hamilton Integrated Research Ethics Board (ID 11240), Centre Hospitalier Universitaire de Québec-Université Laval (37-2021-6561), and Health Research Ethics Board, University of Alberta (Pro00099426), CHU Sainte-Justine, Université de Montréal (MEO-37-2021-3123), Lawson Health Research Institute (ReDA ID 9857), Trillium Health Partners Research Ethics Board (#1002), Queen's University Health Sciences & Affiliated Teaching Hospitals Research Ethics Board (#6029527), IWK Research Ethics Board (#1026214). Newfoundland and Labrador Health Research Ethics Board (# 2020.047)

Classification of chronic health condition was the following: neurological, cardiac, pulmonary (including asthma), metabolic (including diabetes mellitus), renal, and chromosomal disorders; immunosuppression (including malignancy), obesity, prematurity (<37 weeks gestational age), anemia and/or hemoglobinopathy and 'other' comorbidities which included patients who could not be classified into the previous categories.

**eTable 1.** Multivariable logistic regression models focusing only on baseline characteristics as risk factors for severe PCR-positive SARS-CoV-2 infection (WHO COVID-19 clinical progression scale of ≥6). (n=401)
Model 1 incorporates the number of comorbidities, model 2 selected individual comorbidities

| **Covariates** | **Model 1** aOR (95% CI) | **Model 2** aOR (95% CI) |
|---|---|---|
| **Age category** | | |
| <1 year | Ref. | Ref. |
| 1-4 years | 1.41 (0.71-2.79) | 1.45 (0.73-2.86) |
| 5-11 years | 1.52 (0.75-3.05) | 1.32 (0.65-2.68) |
| ≥12 years | **2.22 (1.12-4.40)** | 1.41 (0.66-3.00) |
| **Comorbidities** | | |
| None | Ref. | - |
| 1 comorbidity | 1.32 (0.76-2.29) | - |
| Multiple (≥2) comorbidities | **2.00 (1.09-3.65)** | - |
| Obesity | - | **3.42 (1.66-7.06)** |
| Anemia or hemoglobinopathy | - | **3.14 (1.00-9.88)** |
| Neurological disorders | - | **2.12 (1.13-3.95)** |

aOR adjusted odds ratio; CI confidence intervals

**eTable 2**. Demographic and clinical characteristics of patients PCR-positive SARS-CoV-2 infection (n=403) according to severity of disease (WHO COVID-19 clinical progression scale of ≥6 and/or ICU admission)

| | Mild/Moderate Disease* N=275 (%) | Severe Disease and/or ICU admission* N=128 (%) | p-value** |
|---|---|---|---|
| **Demographics** | | | |
| Median Age in years (IQR)[1] | 3.09 (0.30-10.10) | 6.04 (1.24-12.67) | **0.004** |
| Age ≤30 days | 30 (81.1%) | 7 (18.9%) | 0.115 |
| Age category[1] | | | |
| <1 year | 88 (77.9%) | 25 (22.1%) | **0.024** |
| 1-4 years | 74 (69.8%) | 32 (30.2%) | |
| 5-11 years | 55 (61.8%) | 34 (38.2%) | |
| ≥12 years | 56 (60.2%) | 37 (39.8%) | |
| Male sex | 157 (69.2%) | 70 (30.8%) | 0.730 |
| Admission period | | | |
| Prior to July 1, 2020 | 14 (73.7% | 5 (26.3%) | 0.438 |
| July 1, 2020 to Dec 31, 2020 | 108 (71.5%) | 43 (28.5%) | |
| Jan 1, 2021 to May 31, 2021 | 153 (65.7%) | 80 (34.3%) | |
| Admitted in Canada | 175 (65.8%) | 91 (34.2%) | 0.174 |
| **Number of Comorbidities** | | | |
| None | 157 (72.7%) | 59 (27.3%) | 0.051 |
| At least 1 comorbidity | 118 (63.1%) | 69 (36.9%) | |
| Multiple (≥2) comorbidities | 42 (56.0%) | 33 (44.0%) | **0.017** |
| None | 157 (72.7%) | 59 (27.3%) | **0.028** |
| 1 comorbidity | 76 (67.8%) | 36 (32.1%) | |
| Multiple (≥2) comorbidities | 42 (56.0%) | 33 (44.0%) | |
| **Individual comorbidities**[2] | | | |
| Obesity | 24 (48.0%) | 26 (52.0%) | **0.002** |
| Pulmonary disorders | 32 (59.2%) | 22 (40.7%) | 0.172 |
| Non-asthma pulmonary disorders[3] | 10 (45.4%) | 12 (54.5%) | **0.034** |
| Asthma | 22 (68.7%) | 10 (31.2%) | 1.0 |
| Prematurity (<37 weeks gestational age) | 17 (56.7%) | 13 (43.3%) | 0.226 |
| Immunosuppression | 26 (83.9%) | 5 (16.1%) | 0.069 |
| Malignancy[4] | 15 (93.7%) | 1 (6.2%) | **0.027** |
| Anemia or hemoglobinopathy[5] | 7 (53.8%) | 6 (43.1%) | 0.406 |
| Sickle cell anemia | 4 (66.7%) | 2 (33.3%) | 1.0 |
| Neurological disorders[6] | 30 (53.6%) | 26 (46.4%) | **0.017** |
| Cardiac disorders | 12 (63.1%) | 7 (36.8%) | 0.814 |
| Metabolic disorders | 6 (50.0%) | 6 (50.0%) | 0.288 |
| Diabetes mellitus | 2 (40.0%) | 3 (60.0%) | 0.332 |
| Renal disorders | 7 (50.0%) | 7 (50.0%) | 0.230 |
| Hypertension | 4 (44.4%) | 5 (55.5%) | 0.150 |
| Chromosomal disorders[7] | 3 (37.5%) | 5 (62.5%) | 0.116 |
| Other[8] | 13 (52.0%) | 12 (48.0%) | 0.114 |
| **Clinical Features** | | | |
| Fever prior to or during admission[9] | 207 (68.5%) | 95 (31.4%) | 0.445 |

| | | | |
|---|---|---|---|
| Cough | 135 (64.9%) | 73 (35.1%) | 0.168 |
| Shortness of breath | 100 (53.2%) | 88 (46.8%) | **<0.001** |
| Vomiting | 83 (65.3%) | 44 (34.6%) | 0.466 |
| Rhinitis | 90 (72.0%) | 35 (28.0%) | 0.331 |
| Diarrhea | 81 (70.4%) | 34 (29.6%) | 0.631 |
| Abdominal pain | 70 (64.8%) | 38 (35.2%) | 0.440 |
| Headache | 46 (63.9%) | 26 (36.1%) | 0.462 |
| Rash | 45 (65.2%) | 24 (34.8%) | 0.653 |
| Conjunctivitis | 39 (57.3%) | 29 (42.6%) | **0.049** |
| Myalgia | 40 (66.7%) | 20 (33.3%) | 0.894 |
| Pharyngitis | 46 (76.7%) | 14 (23.3%) | 0.171 |
| Wheeze | 34 (63.0%) | 20 (37.0%) | 0.461 |
| Chest pain | 26 (66.7%) | 13 (33.3%) | 0.967 |
| Cracked lips | 22 (57.9%) | 16 (42.1%) | 0.209 |
| Edema of hands and feet | 16 (59.2%) | 11 (40.7%) | 0.410 |
| Anosmia | 8 (66.7%) | 4 (33.3%) | 1.0 |
| Seizures | 5 (45.4%) | 6 (54.5%) | 0.111 |
| **Coinfections** | | | |
| Bacterial (positive cultures)[10] | 13 (54.2%) | 11 (45.8%) | 0.193 |
| Viral | 15 (46.9%) | 17 (53.1%) | **0.012** |
| Bacterial and/or viral | 28 (51.8%) | 26 (48.1%) | **0.009** |
| **Imaging** | | | |
| No chest imaging performed | 86 (94.5%) | 5 (5.5%) | **<0.001** |
| CXR and/or CT chest normal or had findings unrelated to COVID-19 | 96 (79.3%) | 25 (20.7%) | |
| CXR and/or CT chest abnormalities compatible with COVID-19 | 93 (48.7%) | 98 (51.3%) | |
| **Laboratory investigations** | | | |
| Leukopenia (<4 X$10^9$/L)[11] | 44 (74.6%) | 15 (25.4%) | 0.239 |
| Leukocytosis (>15x$10^9$/L)[11] | 30 (63.8%) | 17 (36.2%) | 0.731 |
| Neutropenia (<1.5x $10^9$/L)[12] | 68 (80.9%) | 16 (19.0%) | **<0.001** |
| Normal neutrophil count[12] | 131 (72.0%) | 51 (28.0%) | |
| Neutrophilia[12] | 56 (48.3%) | 60 (51.7%) | |
| Thrombocytopenia (<100 x $10^9$/L)[13] | 22 (52.4%) | 20 (47.6%) | **0.004** |
| Normal platelet count[13] | 189 (72.4%) | 72 (27.6%) | |
| Thrombocytosis (>450 x $10^9$/L) [13] | 47 (57.3%) | 35 (42.7%) | |
| CRP not done[14] | 49 (81.7%) | 11 (18.3%) | **<0.001** |
| Normal CRP[14] | 165 (73.7%) | 59 (26.3%) | |
| CRP >50 (mg/L)[14] | 61 (51.3%) | 58 (48.7%) | |
| Ferritin >500 (mcg/L)[15] | 24 (43.6%) | 31 (56.4%) | 0.160 |
| Albumin<29 (g/L)[16] | 17 (36.9%) | 29 (63.0%) | **<0.001** |
| **Complications** | | | |
| MIS-C diagnosis | 44 (54.3%) | 37 (45.7%) | **0.004** |

CT computed tomography; CRP C-reactive protein; CXR chest x-ray; IQR interquartile range; MIS-C multisystem inflammatory syndrome in children

* Percentages were calculated for the row, i.e., % of a stratum with severe or mild/moderate disease.

** Comparisons performed using chi squared or Fisher's exact test as appropriate for categorical data and Wilcoxon rank sum test for continuous data as appropriate.

[1] A total of 2 children with mild/moderate disease are missing date of birth.

[2] Children are listed more than once if they had more than one comorbidity.

[3] Clinical information is available for 14/22 patients: obstructive sleep apnea (n=5); bronchopulmonary dysplasia (n=3); pulmonary hypertension (n=2); cystic fibrosis (n=1); chronic aspiration pneumonia (n=1); subglottic stenosis (n=1); Pierre-Robin anomaly (n=1)

[4] Malignancy consists of the following: acute lymphoblastic leukemia (n=11); neuroblastoma (n=2); lymphoma (n=1); rhabdomyosarcoma (n=1); and osteosarcoma (n=1)

[5] Anemia and/or hemoglobinopathy consist of the following: sickle cell disease (n=6); Glucose-6-phosphate dehydrogenase deficiency (n=2); anemia of chronic disease (n=2); ovalocytosis (n=1); pancytopenia of unknown etiology (n=1); iron deficient anemia (n=1)

[6] Additional information is available for 47/56 patients: syndromic condition with global developmental delay (GDD) (n=19); autism and/or GDD of unexplained etiology (n=10); cerebral palsy (n=7); muscular dystrophy (n=3); hydrocephalus (n=2); congenital microcephaly (n=2); brain tumor (n=1); focal cortical dysplasia (n=1); encephalopathy (n=1); epilepsy (n=1)

[7] Additional information is available for 7/8 patients: Trisomy 21 (n=3); unbalanced translocation (n=2); Prader-Willi syndrome (n=1); Miller-Dieker syndrome (n=1)

[8] Other consists of the following: gastrostomy (n=12); liver transplantation (n=2); celiac disease (n=1); Crohn's disease (n=1) failure to thrive (n=1); inflammatory bowel disease (n=1); inflammatory bowel disease and Hirschsprung disease (n=1); unclear liver disease (n=1); esophageal atresia (n=1); scoliosis (n=2); liver cirrhosis & cholestasis (n=1); Glanzmann thrombasthenia (n=1)

[9] A total of 35 children were missing data on fever prior to or during admission (19 with mild/moderate disease and 16 with severe disease) resulting in a total N of 368.

[10] Children with one of the following are included: blood culture thought to be truly positive and/or positive endotracheal tube cultures treated with antibiotics for presumptive bacterial pneumonia and/or positive bacterial culture from another site treated with antibiotics.

[11] A total of 17 children were missing leukocyte values (16 with mild/moderate disease and 1 with severe disease) resulting in a total N of 386.

[12] A total of 21 children were missing neutrophil values (20 with mild/moderate disease and 1 with severe disease) resulting in a total N of 382. Cut-off values for neutrophilia are age dependent.

[13] A total of 18 children were missing a platelet value (17 with mild/moderate disease and 1 with severe disease) resulting in a total N of 385.

[14] A total of 60 children were missing a CRP value (49 with mild/moderate disease and 11 with severe disease) resulting in a total N of 343.

[15] The highest ferritin value measured variable used and a total of 220 children were missing a value (179 with mild/moderate disease and 41 with severe disease) resulting in a total N of 183.

[16] A total of 271 children were missing albumin values (199 with mild/moderate disease and 72 with severe disease) resulting in a total N of 132.

**eTable 3** Age stratified univariable logistic regression analysis among children aged younger than 12 years (n=308) and adolescents 12 years of age and older (n=93) for severe PCR-positive SARS-CoV-2 infection (WHO COVID-19 clinical progression scale of ≥6)

| | **Unadjusted OR among those younger than 12 years of age N=308** | **Unadjusted OR among those 12 years of age and older N=93** |
|---|---|---|
| | **OR (95% CI)** | **OR (95% CI)** |
| **Number of comorbidities** | | |
| None | Ref. | Ref. |
| 1 comorbidity | 1.14 (0.58-2.25) | 1.78 (0.65-4.87) |
| Multiple (≥2) comorbidities | **2.85 (1.45-5.58)** | 1.22 (0.39-3.86) |
| **Individual comorbidities** | | |
| Obesity | **3.70 (1.15-11.86)** | **2.93 (1.22-7.05)** |
| Pulmonary disorders | **2.38 (1.17-4.82)** | 0.69 (0.20-2.40) |
| Non-asthma pulmonary disorders | **4.33 (1.60-11.70)** | 0.44 (0.05-4.08) |
| Anemia or hemoglobinopathy | 3.03 (0.90-10.26) | 1.84 (0.11-30.47) |
| Neurological disorders | **2.74 (1.33-5.63)** | 1.42 (0.51-3.99) |
| Cardiac disorders | 2.43 (0.83-7.09) | - |
| Hypertension | 3.57 (0.70-18.13) | 3.81 (0.33-43.65) |
| Chromosomal disorders | **4.84 (1.06-22.18)** | **-** |
| **Clinical Features** | | |
| Fever prior to or during admission | 0.86 (0.42-1.77) | 4.05 (0.84-19.44) |
| Shortness of breath | **5.98 (3.30-10.86)** | **3.73 (1.26-11.02)** |
| Conjunctivitis | 1.63 (0.85-3.10) | 1.61 (0.45-5.73) |
| Pharyngitis | 0.40 (0.13-1.17) | 0.52 (0.18-1.47) |
| Wheeze | 1.86 (0.94-3.70) | 0.71 (0.13-3.88) |
| Seizures | 2.88 (0.75-11.03) | 1.84 (0.11-30.47) |
| **Coinfections** | | |
| Bacterial and/or viral | **4.11 (2.10-8.02)** | 1.24 (0.32-4.76) |
| **Imaging** | | |
| CXR and/or CT normal or had findings unrelated to COVID-19 | Ref. | Ref. |
| No chest imaging done | **0.07 (0.01-0.52)** | 0.54 (0.04-6.77) |
| CXR or CT abnormalities compatible with COVID-19 | **3.57 (1.90-6.70)** | **5.57 (1.16-26.69)** |
| **Laboratory investigations** | | |
| Neutropenia (<1.5x $10^9$/L) | 0.72 (0.32-1.66) | 1.38 (0.42-4.52) |
| Normal neutrophil count | Ref. | Ref. |
| Neutrophilia | **3.02 (1.64-5.58)** | **3.23 (1.17-8.94)** |
| Thrombocytopenia (<100 x $10^9$/L) | 1.79 (0.78-4.07) | 1.10 (0.24-5.01) |
| Normal platelet count | Ref. | Ref. |
| Thrombocytosis (>450 x $10^9$/L) | 1.66 (0.89-3.09) | 2.30 (0.56-9.35) |
| CRP not done | Ref. | Ref. |
| Normal CRP | 1.18 (0.53-2.65) | - |
| CRP >50 (mg/L) | 2.04 (0.86-4.84) | - |
| **Complications** | | |
| MIS-C | 1.52 (0.81-2.85) | 1.60 (0.56-4.55) |

Note. Among those less than 12 years of age, N for model with fever is 281 as 27 deleted due to missingness, N for models with neutrophil counts are 292 as 16 deleted due to missingness, N for models with platelet counts are 295 as 13 deleted due to missingness. Among those 12 years of age and older, N for model with fever is 86 as 7 deleted due to missingness, N for models with neutrophil counts and platelet counts are 88 as 5 deleted due to missingness.

OR odds ratio; CT computed tomography; CRP C-reactive protein; CXR chest x-ray; IQR interquartile range; MIS-C multisystem inflammatory syndrome in children

**eTable 4.** Demographic and clinical characteristics among children aged less than 12 years (n=308) according to according to severity of disease (WHO COVID-19 clinical progression scale of ≥6)

| | **Mild/Moderate Disease*** **N=239 (%)** | **Severe Disease*** **N=69 (%)** | **p-value**** |
|---|---|---|---|
| **Demographics** | | | |
| Male sex | 136 (76.8%) | 41 (23.2%) | 0.815 |
| Admission period | | | |
| Prior to July 1, 2020 | 13 (81.2%) | 3 (18.7%) | 0.973 |
| Between July 1, 2020 to Dec 31, 2020 | 97 (78.2%) | 27 (21.8%) | |
| Between Jan 1, 2021 to May 31, 2021 | 129 (76.8%) | 39 (23.2%) | |
| Admitted in Canada | 138 (77.5%) | 40 (22.5%) | 1.0 |
| **Number of Comorbidities** | | | |
| None | 150 (81.5%) | 34 (18.5%) | 0.061 |
| At least 1 comorbidity | 89 (71.8%) | 35 (28.2%) | |
| Multiple (≥2) comorbidities | 31 (60.8%) | 20 (39.2%) | **0.001** |
| None | 150 (81.5%) | 34 (18.5%) | **0.007** |
| 1 comorbidity | 58 (79.4%) | 15 (20.5%) | |
| Multiple (≥2) comorbidities | 31 (60.8%) | 20 (39.2%) | |
| **Individual comorbidities**[1] | | | |
| Obesity | 6 (50.0%) | 6 (50.0%) | **0.047** |
| Pulmonary disorders | 25 (62.5%) | 15 (37.5%) | **0.024** |
| Non-asthma pulmonary disorders | 8 (47.0%) | 9 (52.9%) | **0.005** |
| Asthma | 17 (73.9%) | 6 (26.1%) | 0.857 |
| Prematurity (<37 weeks gestational age) | 20 (71.4%) | 8 (28.6%) | 0.560 |
| Immunosuppression | 18 (81.8%) | 4 (18.2%) | 0.793 |
| Cancer | 12 (92.3%) | 1 (7.7%) | 0.310 |
| Anemia or hemoglobinopathy | 6 (54.5%) | 5 (45.4%) | 0.073 |
| Sickle cell anemia | 3 (75.0%) | 1 (25.0%) | 1.0 |
| Neurological disorders | 22 (59.4%) | 15 (40.5%) | **0.009** |
| Cardiac disorders | 9 (60.0%) | 6 (40.0%) | 0.174 |
| Metabolic disorders | 5 (83.3%) | 1 (16.7%) | 1.0 |
| Diabetes mellitus | 1 (100.0%) | 0 | 1.0 |
| Renal disorders | 7 (58.3%) | 5 (41.7%) | 0.201 |
| Hypertension | 3 (50.0%) | 3 (50.0%) | 0.128 |
| Chromosomal disorders | 3 (42.8%) | 4 (57.1%) | **0.047** |
| Other | 11 (64.7%) | 6 (35.3%) | 0.311 |
| **Clinical Features** | | | |
| Fever prior to or during admission[2] | 182 (79.5%) | 47 (20.5%) | 0.826 |
| Cough | 104 (75.4%) | 34 (24.6%) | 0.478 |
| Shortness of breath | 73 (59.3%) | 50 (40.6%) | **<0.001** |
| Vomiting | 67 (72.8%) | 25 (27.2%) | 0.245 |
| Rhinitis | 84 (77.8%) | 24 (22.2%) | 1.0 |
| Diarrhea | 64 (77.1%) | 19 (22.9%) | 1.0 |
| Abdominal pain | 61 (76.2%) | 19 (23.7%) | 0.857 |
| Headache | 33 (75.0%) | 11 (25.0%) | 0.802 |

| | | | |
|---|---|---|---|
| Rash | 44 (74.6%) | 15 (25.4%) | 0.656 |
| Conjunctivitis | 40 (70.2%) | 17 (29.8%) | 0.189 |
| Myalgia | 31 (79.5%) | 8 (20.5%) | 0.922 |
| Pharyngitis | 32 (88.9%) | 4 (11.1%) | 0.092 |
| Wheeze | 31 (67.4%) | 15 (32.6%) | 0.108 |
| Chest pain | 11 (73.3%) | 4 (26.7%) | 0.751 |
| Cracked lips | 24 (72.7%) | 9 (27.3%) | 0.625 |
| Edema of hands and feet | 16 (72.7%) | 6 (27.3%) | 0.762 |
| Anosmia | 4 (100.0%) | 0 | 0.578 |
| Seizures | 5 (55.5%) | 4 (44.4%) | 0.117 |
| **Coinfections** | | | |
| Bacterial (positive cultures)[3] | 10 (55.5%) | 8 (44.4%) | **0.043** |
| Viral | 13 (46.4%) | 15 (53.6%) | **<0.001** |
| Bacterial and/or viral | 23 (52.3%) | 21 (47.7%) | **<0.001** |
| **Imaging** | | | |
| No chest imaging done | 77 (98.7%) | 1 (1.3%) | |
| CXR and/or CT chest normal or had findings unrelated to COVID-19 | 88 (83.8%) | 17 (16.2%) | **<0.001** |
| CXR and/or CT chest abnormalities compatible with COVID-19 | 74 (59.2%) | 51 (40.8%) | |
| **Laboratory investigations** | | | |
| Leukopenia (<4 X$10^9$/L)[4] | 33 (91.7%) | 3 (8.3%) | **0.033** |
| Leukocytosis (>15x$10^9$/L)[4] | 30 (71.4%) | 12 (28.6%) | 0.472 |
| Neutropenia (<1.5x $10^9$/L) [5] | 58 (86.6%) | 9 (13.4%) | |
| Normal neutrophil count[5] | 112 (82.3%) | 24 (17.6%) | **<0.001** |
| Neutrophilia[5] | 54 (60.7%) | 35 (39.3%) | |
| Thrombocytopenia (<100 x $10^9$/L)[6] | 23 (69.7%) | 10 (30.3%) | |
| Normal platelet count[6] | 152 (80.4 %) | 37 (19.6%) | 0.164 |
| Thrombocytosis (>450 x $10^9$/L) [6] | 52 (71.2%) | 21 (28.8%) | |
| CRP not done[7] | 42 (82.3%) | 9 (17.6%) | |
| Normal CRP[7] | 142 (79.8%) | 36 (20.2%) | 0.133 |
| CRP >50 (mg/L)[7] | 55 (69.6%) | 24 (30.4%) | |
| Ferritin >500 (mcg/L)[8] | 24 (63.1%) | 14 (36.8%) | 0.523 |
| Albumin<29 (g/L)[9] | 21 (56.7%) | 16 (43.2%) | **0.033** |
| **Complications** | | | |
| MIS-C diagnosis | 45 (71.4%) | 18 (28.6%) | 0.251 |

CT computed tomography; CRP C-reactive protein; CXR chest x-ray; MIS-C multisystem inflammatory syndrome in children

* Percentages were calculated for the row, i.e., % of a stratum with severe or mild/moderate disease.

** Comparisons performed using chi squared or Fisher's exact test as appropriate for categorical data.

[1] Children are listed more than once if they had more than one comorbidity.

[2] A total of 27 children were missing data on fever prior to or during admission (17 with mild/moderate disease and 10 with severe disease) resulting in a total N of 281.

[3] Children with one of the following are included: blood culture thought to be truly positive and/or positive endotracheal tube cultures treated with antibiotics for presumptive bacterial pneumonia and/or positive bacterial culture from another site treated with antibiotics.

[4] A total of 13 children were missing leukocyte values (12 with mild/moderate disease and 1 with severe disease) resulting in a total N of 295.

[5] A total of 16 children were missing neutrophil values (15 with mild/moderate disease and 1 with severe disease) resulting in a total N of 292. Cut-off values for neutrophilia are age dependent

[6] A total of 13 children were missing platelet values (12 with mild/moderate disease and 1 with severe disease) resulting in a total N of 295.

[7] A total of 51 children were missing a CRP value (42 with mild/moderate disease and 9 with severe disease) resulting in a total N of 257.

[8] The highest ferritin value measured variable used and a total of 181 children were missing a value (152 with mild/moderate disease and 29 with severe disease) resulting in a total N of 127.

[9] A total of 205 children were missing albumin values (166 with mild/moderate disease and 39 with severe disease) resulting in a total N of 103.

**eTable 5.** Demographic and clinical characteristics among adolescents aged 12 to 17 years (n=93) according severity of disease (WHO COVID-19 clinical progression scale of ≥6)

| | Mild/Moderate Disease* N=60 (%) | Severe Disease* N=33 (%) | p-value** |
|---|---|---|---|
| **Demographics** | | | |
| Male sex | 34 (69.4%) | 15 (30.6%) | 0.413 |
| Admission period | | | |
| Prior to July 1, 2020 | 3 (100.0%) | 0 | |
| Between July 1, 2020 to Dec 31, 2020 | 20 (76.9%) | 6 (23.1%) | 0.112 |
| Between Jan 1, 2021 to May 31, 2021 | 37 (57.8%) | 27 (42.2%) | |
| Admitted in Canada | 56 (64.4%) | 31 (35.6%) | 1.0 |
| **Number of comorbidities** | | | |
| None | 22 (71.0%) | 9 (29.0%) | 0.490 |
| At least 1 comorbidity | 38 (61.3%) | 24 (38.7%) | |
| Multiple (≥2) comorbidities | 16 (66.7%) | 8 (33.3%) | 0.994 |
| None | 22 (71.0%) | 9 (29.0%) | |
| 1 comorbidity | 22 (57.9%) | 16 (42.1%) | 0.512 |
| Multiple (≥2) comorbidities | 16 (66.7%) | 8 (33.3%) | |
| **Individual comorbidities[1]** | | | |
| Obesity | 19 (50.0%) | 19 (50.0%) | **0.027** |
| Pulmonary disorders | 10 (71.4%) | 4 (28.6%) | 0.763 |
| Non-asthma pulmonary disorders | 4 (80.0%) | 1 (20.0%) | 0.652 |
| Asthma | 6 (66.7%) | 3 (33.3%) | 1.0 |
| Prematurity (<37 weeks gestational age) | 0 | 2 (100.0%) | 0.123 |
| Immunosuppression | 7 (87.5%) | 1 (12.5%) | 0.252 |
| Cancer | 2 (100.0%) | 0 | 0.537 |
| Anemia or hemoglobinopathy | 1 (50.0%) | 1 (50.0%) | 1.0 |
| Sickle cell anemia | 1 (50.0%) | 1 (50.0%) | 1.0 |
| Neurological disorders | 11 (57.9%) | 8 (42.1%) | 0.684 |
| Cardiac disorders | 4 (100.0%) | 0 | 0.293 |
| Metabolic disorders | 3 (50.0%) | 3 (50.0%) | 0.662 |
| Diabetes mellitus | 2 (50.0%) | 2 (50.0%) | 0.613 |
| Renal disorders | 2 (100.0%) | 0 | 0.537 |
| Hypertension | 1 (33.3%) | 2 (66.7%) | 0.286 |
| Chromosomal disorders | 0 | 1 (100.0%) | 0.355 |
| Other | 6 (75.0%) | 2 (25.0%) | 0.707 |
| **Clinical Features** | | | |
| Fever prior to or during admission[2] | 43 (59.7%) | 29 (40.3%) | 0.075 |
| Cough | 41 (60.3%) | 27 (39.7%) | 0.246 |
| Shortness of breath | 36 (56.2%) | 28 (43.7%) | **0.019** |
| Vomiting | 23 (67.6%) | 11 (32.3%) | 0.799 |
| Rhinitis | 12 (75.0%) | 4 (25.0%) | 0.401 |
| Diarrhea | 23 (71.9%) | 9 (28.1%) | 0.397 |
| Abdominal pain | 19 (67.8%) | 9 (32.1%) | 0.837 |
| Headache | 19 (67.8%) | 9 (32.1%) | 0.837 |
| Rash | 6 (60.0%) | 4 (40.0%) | 0.739 |
| Conjunctivitis | 6 (54.5%) | 5 (45.4%) | 0.512 |

| | | | |
|---|---|---|---|
| Myalgia | 15 (71.4%) | 6 (28.6%) | 0.622 |
| Pharyngitis | 18 (75.0%) | 6 (25.0%) | 0.318 |
| Wheeze | 5 (71.4%) | 2 (28.6%) | 1.0 |
| Chest pain | 16 (66.7%) | 8 (33.3%) | 0.994 |
| Cracked lips | 2 (40.0%) | 3 (60.0%) | 0.343 |
| Edema of hands and feet | 3 (60.0%) | 2 (40.0%) | 1.0 |
| Anosmia | 6 (75.0%) | 2 (25.0%) | 0.707 |
| Seizures | 1 (50.0%) | 1 (50.0%) | 1.0 |
| **Coinfections** | | | |
| Bacterial (positive cultures)[3] | 4 (66.7%) | 2 (33.3%) | 1.0 |
| Viral | 2 (50.0%) | 2 (50.0%) | 0.613 |
| Bacterial and/or viral | 6 (60.0%) | 4 (40.0%) | 0.739 |
| **Imaging** | | | |
| No chest imaging done | 12 (92.3%) | 1 (7.7%) | **0.0039** |
| CXR and/or CT chest normal or had findings unrelated to COVID-19 | 13 (86.7%) | 2 (13.3%) | |
| CXR and/or CT chest abnormalities compatible with COVID-19 | 35 (53.8%) | 30 (46.1%) | |
| **Laboratory investigations** | | | |
| Leukopenia (<4 X$10^9$/L)[4] | 14 (60.9%) | 9 (39.1%) | 1.0 |
| Leukocytosis (>15x$10^9$/L)[4] | 1 (33.3%) | 2 (66.7%) | 0.552 |
| Neutropenia (<1.5x $10^9$/L)[5] | 11 (64.7%) | 6 (35.3%) | 0.068 |
| Normal neutrophil count[5] | 33 (71.7%) | 13 (28.3%) | |
| Neutrophilia[5] | 11 (44.0%) | 14 (56.0%) | |
| Thrombocytopenia (<100 x $10^9$/L)[6] | 5 (62.5%) | 3 (37.5%) | 0.505 |
| Normal platelet count[6] | 46 (64.8%) | 25 (35.2%) | |
| Thrombocytosis (>450 x $10^9$/L)[6] | 4 (44.4%) | 5 (55.5%) | |
| CRP not done[7] | 9 (100.0%) | 0 | **0.0147** |
| Normal CRP[7] | 30 (68.2%) | 14 (31.8%) | |
| CRP >50 (mg/L)[7] | 21 (52.5%) | 19 (47.5%) | |
| Ferritin >500 (mcg/L)[8] | 6 (35.3%) | 11 (64.7%) | 0.281 |
| Albumin<29 (g/L)[9] | 3 (33.3%) | 6 (66.7%) | 0.106 |
| **Complications** | | | |
| MIS-C diagnosis | 10 (55.5%) | 8 (44.4%) | 0.541 |

CT computed tomography; CRP C-reactive protein; CXR chest x-ray; MIS-C multisystem inflammatory syndrome in children; PCR polymerase chain reaction; WHO World Health Organization.

* Percentages were calculated for the row, i.e., % of a stratum with severe or mild/moderate disease.

** Comparisons performed using chi squared or Fisher's exact test as appropriate for categorical data.

[1] Children are listed more than once if they had more than one comorbidity.

[2] A total of 7 children were missing data on fever prior to or during admission (5 with mild/moderate disease and 2 with severe disease) resulting in a total N of 86.

[3] Children with one of the following are included: blood culture thought to be truly positive and/or positive endotracheal tube cultures treated with antibiotics for presumptive bacterial pneumonia and/or positive bacterial culture from another site treated with antibiotics.

[4] A total of 4 children were missing leukocyte values (4 with mild/moderate disease) resulting in a total N of 89.

[5] A total of 5 children were missing neutrophil values (5 with mild/moderate disease) resulting in a total N of 88. Cut-off values for neutrophilia are age dependent

[6] A total of 5 children were missing platelet values (5 with mild/moderate disease) resulting in a total N of 88.

[7] A total of 9 children were missing a CRP value (9 with mild/moderate disease) resulting in a total N of 84.

[8] The highest ferritin value measured variable used and a total of 38 children were missing a value (33 with mild/moderate disease and 35 with severe disease) resulting in a total N of 55.

[9] A total of 64 children were missing albumin values (43 with mild/moderate disease and 21 with severe disease) resulting in a total N of 29.

**eTable 6.** Multivariable logistic regression models for factors associated with severe PCR-positive SARS-CoV-2 infection (WHO COVID-19 progression scale of ≥6), stratified according to age groups.

Model 1 incorporates selected individual comorbidities, model 2 the number of comorbidities while excluding individual comorbidities; both in children <12 years of age.

Model 3 is incorporates selected individual comorbidities in adolescents ≥ 12 years

| **Covariates** | **Model 1: Children < 12 years old** aOR (95% CI) | **Model 2: Children < 12 years old** aOR (95% CI) | **Model 3: Children ≥ 12 years old** aOR (95% CI) |
|---|---|---|---|
| **Number of comorbidities** | | | |
| None | - | Ref. | - |
| 1 comorbidity | - | 1.57 (0.69-3.60) | - |
| Multiple (>2) comorbidities | - | **2.60 (1.12-6.06)** | - |
| **Individual comorbidities** | | | |
| Obesity | 1.17 (0.29-4.67) | - | **3.21 (1.15-8.93)** |
| Non-asthma pulmonary disorders | 1.68 (-0.45-6.22) | - | - |
| Anemia or hemoglobinopathy | - | - | - |
| Neurological disorders | **3.16 (1.19-8.43)** | - | - |
| Cardiac disorders | - | - | - |
| Hypertension | - | - | - |
| Chromosomal disorders | - | - | - |
| **Coinfections** | | | |
| Bacterial and/or viral | **2.60 (1.13-5.99)** | **2.50 (1.10-5.69)** | - |
| **Imaging** | | | |
| No chest imaging done | 0.15 (0.02-1.24) | 0.16 (0.02-1.28) | 0.82 (0.05-8.93) |
| CXR and/or CT normal or had findings unrelated to COVID-19 | Ref. | Ref. | Ref. |
| CXR or CT abnormalities compatible with COVID-19 | **3.10 (1.49-6.42)** | **2.99 (1.45-6.18)** | **7.87 (1.32-46.97)** |
| **Clinical presentation** | | | |
| Shortness of breath | **5.02 (2.10-12.04)** | **5.03 (2.12-11.88)** | - |
| **Complications** | | | |
| MIS-C diagnosis | **4.85 (1.70-13.83)** | **4.92 (1.72-14.04)** | 1.11 (0.27-4.67) |
| **Laboratory investigations** | | | |
| Neutrophilia | 1.54 (0.74-3.22) | 1.55 (0.75-3.22) | **5.81 (1.85-18.22)** |

aOR adjusted odds ratio; CI confidence interval; CT computed tomography; CXR chest x-ray; MIS-C multisystem inflammatory syndrome in children

**eTable 7.** Demographic and clinical characteristics among non-MIS-C patients (n=322) with PCR-positive SARS-CoV-2 infection, according to severity of disease (WHO COVID-19 clinical progression scale of ≥6)

| | **Mild/Moderate Disease* N=246 (%)** | **Severe Disease* N=76 (%)** | **p-value**** | **Unadjusted OR for severe disease** |
|---|---|---|---|---|
| **Demographics** | | | | |
| Median Age in years (IQR)[1] | 2.08 (0.23-9.66) | 3.65 (0.98-14.41) | **0.0062** | **1.04 (1.00-1.09)** |
| Age ≤30 days | 34 (91.9%) | 3 (8.1%) | **0.0218** | **0.26 (0.08-0.86)** |
| Age category[1] | | | | |
| <1 year | 94 (83.2%) | 19 (16.8%) | **0.0303** | Ref. |
| 1-4 years | 61 (71.8%) | 24 (28.2%) | | 1.95 (0.98-3.85) |
| 5-11 years | 39 (83.0%) | 8 (17.0%) | | 1.01 (0.41-2.51) |
| ≥12 years | 50 (66.7%) | 25 (33.3%) | | **2.47 (1.24-4.92)** |
| Male sex | 137 (76.5%) | 42 (23.5%) | 1.0 | 0.98 (0.58-1.65) |
| Admission period | | | | |
| Prior to July 1, 2020 | 15 (88.2%) | 2 (11.8%) | 0.280 | Ref. |
| Between July 1, 2020 to Dec 31, 2020 | 93 (79.5%) | 24 (20.5%) | | 1.93 (0.41-9.05) |
| Between Jan 1, 2021 to May 31, 2021 | 138 (73.4%) | 50 (26.6%) | | 2.72 (0.60-12.31) |
| Admitted in Canada | 158 (76.3%) | 49 (23.7%) | 1.0 | 1.01 (0.59-1.73) |
| **Number of comorbidities** | | | | |
| None | 128 (83.7%) | 25 (16.3%) | **0.0053** | Ref. |
| At least 1 comorbidity | 118 (69.8%) | 51 (30.2%) | | **2.21 (1.29-3.80)** |
| Multiple (≥2) comorbidities | 44 (63.8%) | 25 (36.2%) | **0.0086** | **2.25 (1.26-4.01)** |
| None | 128 (83.7%) | 25 (16.3%) | **0.0043** | Ref. |
| 1 comorbidity | 74 (74.0%) | 26 (26.0%) | | 1.80 (0.97-3.34) |
| Multiple (≥2) comorbidities | 44 (63.8%) | 25 (36.2%) | | **2.91 (1.52-5.58)** |
| **Individual comorbidities[2]** | | | | |
| Obesity | 24 (55.8%) | 19 (44.2%) | **0.0013** | **3.08 (1.58-6.02)** |
| Pulmonary disorders | 32 (64.0%) | 18 (36.0%) | **0.0389** | **2.07 (1.09-3.96)** |
| Non-asthma pulmonary disorders | 12 (54.5%) | 10 (45.4%) | **0.0250** | **2.95 (1.22-7.14)** |
| Asthma | 20 (71.4%) | 8 (28.6%) | 0.678 | 1.33 (0.56-3.15) |
| Prematurity (<37 weeks gestational age) | 19 (65.5%) | 10 (34.5%) | 0.223 | 1.81 (0.80-4.08) |
| Immunosuppression | 24 (85.7%) | 4 (14.3%) | 0.350 | 0.51 (0.17-1.53) |
| Cancer | 13 (92.8%) | 1 (7.1%) | 0.201 | 0.24 (0.03-1.86) |
| Anemia or hemoglobinopathy | 7 (63.6%) | 4 (36.4%) | 0.295 | 1.90 (0.54-6.66) |
| Sickle cell anemia | 4 (80.0%) | 1 (20.0%) | 1.0 | 0.81 (0.09-7.33) |
| Neurological disorders | 29 (56.9%) | 22 (43.1%) | **<0.001** | **3.05 (1.62-5.72)** |
| Cardiac disorders | 13 (68.4%) | 6 (31.6%) | 0.572 | 1.54 (0.56-4.19) |
| Metabolic disorders | 8 (72.7%) | 3 (27.3%) | 0.725 | 1.22 (0.32-4.73) |
| Diabetes mellitus | 3 (75.0%) | 1 (25.0%) | 1.0 | 1.08 (0.11-10.54) |
| Renal disorders | 9 (69.2%) | 4 (30.8%) | 0.514 | 1.46 (0.44-4.89) |
| Hypertension | 4 (50.0%) | 4 (50.0%) | 0.093 | 3.36 (0.82-13.78) |

| | | | | |
|---|---|---|---|---|
| Chromosomal disorders | 2 (28.6%) | 5 (71.4%) | **0.0092** | **8.59 (1.63-45.23)** |
| Other | 15 (65.2%) | 8 (34.8%) | 0.291 | 1.81 (0.74-4.45) |
| **Clinical Features** | | | | |
| Fever prior to or during admission[3] | 172 (77.5%) | 50 (22.5%) | 0.955 | 1.08 (0.55-2.11) |
| Cough | 132 (71.7%) | 52 (28.3%) | **0.0323** | **1.87 (1.08-3.23)** |
| Shortness of breath | 105 (61.0%) | 67 (38.9%) | **<0.001** | **10.00 (4.77-20.96)** |
| Vomiting | 56 (80.0%) | 14 (20.0%) | 0.520 | 0.77 (0.40-1.47) |
| Rhinitis | 85 (78.0%) | 24 (22.0%) | 0.734 | 0.87 (0.50-1.51) |
| Diarrhea | 57 (82.6%) | 12 (17.4%) | 0.226 | 0.62 (0.31-1.23) |
| Abdominal pain | 35 (81.4%) | 8 (18.6%) | 0.525 | 0.71 (0.31-1.60) |
| Headache | 26 (78.8%) | 7 (21.2%) | 0.900 | 0.86 (0.36-2.06) |
| Rash | 15 (78.9%) | 4 (21.0%) | 1.0 | 0.85 (0.27-2.66) |
| Conjunctivitis | 7 (87.5%) | 1 (12.5%) | 0.686 | 0.45 (0.05-3.76) |
| Myalgia | 30 (76.9%) | 9 (23.1%) | 1.0 | 0.97 (0.44-2.14) |
| Pharyngitis | 32 (88.9%) | 4 (11.1%) | 0.063 | 0.37 (0.13-1.09) |
| Wheeze | 36 (69.2%) | 16 (30.8%) | 0.250 | 1.55 (0.81-2.99) |
| Chest pain | 26 (81.2%) | 6 (18.7%) | 0.644 | 0.72 (0.29-1.83) |
| Cracked lips | 2 (66.7%) | 1 (33.3%) | 0.555 | 1.63 (0.14-18.19) |
| Edema of hands and feet | 1 (50.0%) | 1 (50.0%) | 0.417 | 3.27 (0.20-52.86) |
| Anosmia | 7 (87.5%) | 1 (12.5%) | 0.686 | 0.45 (0.05-3.76) |
| Seizures | 6 (54.5%) | 5 (45.4%) | 0.139 | 2.82 (0.83-9.50) |
| **Coinfections** | | | | |
| Bacterial (positive cultures)[4] | 10 (52.6%) | 9 (47.4%) | **0.0253** | **3.17 (1.24-8.12)** |
| Viral | 13 (46.4%) | 15 (53.6%) | **<0.001** | **4.41 (1.99-9.75)** |
| Bacterial and/or viral | 23 (51.1%) | 22 (48.9%) | **<0.001** | **3.95 (2.05-7.61)** |
| **Imaging** | | | | |
| No chest imaging done | 72 (97.3%) | 2 (2.7%) | **<0.001** | Ref. |
| CXR and/or CT chest normal or had findings unrelated to COVID-19 | 78 (85.7%) | 13 (14.3%) | | **0.17 (0.04-0.76)** |
| CXR and/or CT chest abnormalities compatible with COVID-19 | 96 (61.1%) | 61 (38.8%) | | **3.81 (1.95-7.44)** |
| **Laboratory investigations** | | | | |
| Leukopenia (<4 X10$^9$/L)[5] | 42 (77.8%) | 12 (22.2%) | 0.798 | 0.86 (0.42-1.73) |
| Leukocytosis (>15x10$^9$/L)[5] | 31 (77.5%) | 9 (22.5%) | 0.905 | 0.88 (0.40-1.94) |
| Neutropenia (<1.5x 10$^9$/L)[6] | 64 (81.0%) | 15 (19.0%) | **0.0055** | 0.85 (0.43-1.68) |
| Normal neutrophil count[6] | 124 (78.5%) | 34 (21.5%) | | Ref. |
| Neutrophilia[6] | 39 (60.0%) | 26 (40.0%) | | **2.43 (1.30-4.54)** |
| Thrombocytopenia (<100 x 10$^9$/L)[7] | 19 (73.1%) | 7 (26.9%) | 0.291 | 1.27 (0.51-3.19) |
| Normal platelet count[7] | 176 (77.5%) | 51 (22.5%) | | Ref. |
| Thrombocytosis (>450 x 10$^9$/L)[7] | 35 (67.3%) | 17 (32.7%) | | 1.68 (0.87-3.24) |
| CRP not done[8] | 51 (86.4%) | 8 (13.5%) | **0.0033** | Ref. |
| Normal CRP[8] | 168 (77.4%) | 49 (22.6%) | | 1.86 (0.83-4.18) |
| CRP >50 (mg/L)[8] | 27 (58.7%) | 19 (41.3%) | | **4.49 (1.74-11.58)** |

| | | | | |
|---|---|---|---|---|
| Ferritin >500 (mcg/L)[9] | 10 (52.6%) | 9 (47.4%) | 0.697 | 1.39 (0.51-3.78) |
| Albumin<29 (g/L)[10] | 4 (36.4%) | 7 (63.6%) | **0.0345** | **4.53 (1.17-17.47)** |

OR, odds ratio; CT computed tomography; CRP C-reactive protein; CXR chest x-ray; IQR interquartile range; MIS-C multisystem inflammatory syndrome in children; PCR polymerase chain reaction; WHO World Health Organization.

* Percentages were calculated for the row, i.e., % of a stratum with severe or mild/moderate disease.

** Comparisons performed using chi squared or Fisher's exact test as appropriate for categorical data and Wilcoxon rank sum test for continuous data as appropriate.

[1] A total of 2 children with mild/moderate disease are missing date of birth.

[2] Children are listed more than once if they had more than one comorbidity.

[3] A total of 34 children were missing data on fever prior to or during admission (22 with mild/moderate disease and 12 with severe disease) resulting in a total N of 288.

[4] Children with one of the following are included: blood culture thought to be truly positive and/or positive endotracheal tube cultures treated with antibiotics for presumptive bacterial pneumonia and/or positive bacterial culture from another site treated with antibiotics.

[5] A total of 16 children were missing leukocyte values (15 with mild/moderate disease and 1 with severe disease) resulting in a total N of 306.

[5] A total of 20 children were missing neutrophil values (19 with mild/moderate disease and 1 with severe disease) resulting in a total N of 302. Cut-off values for neutrophilia are age dependent.

[7] A total of 17 children were missing platelet values (16 with mild/moderate disease and 1 with severe disease) resulting in a total N of 305.

[8] A total of 59 children were missing a CRP value (51 with mild/moderate disease and 8 with severe disease) resulting in a total N of 263.

[9] The highest ferritin value measured variable used and a total of 219 children were missing a value (185 with mild/moderate disease and 34 with severe disease) resulting in a total N of 103.

[10] A total of 250 children were missing albumin values (198 with mild/moderate disease and 52 with severe disease) resulting in a total N of 72.

**eTable 8.** Multivariable logistic regression models among non-MIS-C patients for factors associated with severe PCR-positive SARS-CoV-2 infection (WHO COVID-19 clinical progression scale of ≥6).
Model 1 incorporates the number of comorbidities, model 2 selected individual comorbidities.

| Covariates | Model 1<br>aOR (95% CI) | Model 2<br>aOR (95% CI) |
|---|---|---|
| **Age category** | | |
| <1 year | Ref. | Ref. |
| 1-4 years | 1.00 (0.41-2.47) | 1.04 (0.43-2.52) |
| 5-11 years | 0.36 (0.11-1.18) | 0.31 (0.09-1.08) |
| ≥12 years | 0.86 (0.35-2.14) | 0.62 (0.23-1.70) |
| **Number of comorbidities** | | |
| None | Ref. | - |
| 1 comorbidity | 2.04 (0.92-4.54) | - |
| Multiple (≥2) comorbidities | **2.72 (1.16-6.39)** | - |
| **Individual comorbidities** | | |
| Obesity | - | **2.67 (1.00-7.15)** |
| Neurological disorder | - | **4.39 (1.88-10.28)** |
| **Coinfections** | | - |
| Bacterial and/or viral | **2.56 (1.17-5.59)** | **2.84 (1.28-6.30)** |
| **Imaging** | | |
| No chest imaging done | 0.47 (0.09-2.36) | 0.43 (0.08-2.22) |
| CXR and/or CT normal or had findings unrelated to COVID-19 | Ref. | Ref. |
| CXR or CT abnormalities compatible with COVID-19 | **3.05 (1.38-6.71)** | **3.03 (1.36-6.77)** |
| **Clinical presentation** | | |
| Shortness of breath | **5.17 (2.28-11.69)** | **4.92 (2.12-11.41)** |
| **Laboratory investigations** | | |
| Neutrophilia | 1.89 (0.91-3.92) | 1.98 (0.94-4.20) |

aOR adjusted odds ratio; CI confidence interval; CRP C-reactive protein; CT computed tomography; CXR chest x-ray
